# Identification of Parkinson PACE subtypes and repurposing treatments through integrative analyses of multimodal clinical progression, neuroimaging, genetic, and transcriptomic data

**DOI:** 10.1101/2021.07.18.21260731

**Authors:** Chang Su, Yu Hou, Jielin Xu, Zhenxing Xu, Jie Xu, Matthew Brendel, Jacqueline R. M. A. Maasch, Zilong Bai, Haotan Zhang, Yingying Zhu, Molly C. Cincotta, Xinghua Shi, Claire Henchcliffe, James B. Leverenz, Jeffrey Cummings, Michael S. Okun, Jiang Bian, Feixiong Cheng, Fei Wang

## Abstract

Parkinson’s disease (PD) is a progressive neurodegenerative disorder marked by significant clinical and progression heterogeneity resulting from complex pathophysiological mechanisms. This study aimed at addressing heterogeneity of PD through the integrative analysis of a broad spectrum of data sources. We analyzed clinical progression data spanning over 5 years from individuals with *de novo* PD, using machine learning and deep learning, to characterize individuals’ phenotypic progression trajectories for subtyping. We discovered three pace subtypes of PD which exhibited distinct progression patterns and were stable over time: the Inching Pace subtype (PD-I) with mild baseline severity and mild progression speed; the Moderate Pace subtype (PD-M) with mild baseline severity but advancing at a moderate progression rate; and the Rapid Pace subtype (PD-R) with the most rapid symptom progression rate. We found that cerebrospinal fluid P-tau/α-synuclein ratio and atrophy in certain brain regions measured by neuroimaging might be indicative markers of these subtypes. Furthermore, through genetic and transcriptomic data analyses enhanced by network medicine approaches, we detected molecular modules associated with each subtype. For instance, the PD-R-specific module suggested *STAT3*, *FYN*, *BECN1*, *APOA1*, *NEDD4*, and *GATA2* as potential driver genes of PD-R. Pathway analysis suggested that neuroinflammation, oxidative stress, metabolism, AD, PI3K/AKT, and angiogenesis pathways may drive rapid PD progression (i.e., PD-R). Moreover, we identified candidate repurposable drugs via targeting these subtype-specific molecular modules and estimated their treatment effects using two large-scale real-world patient databases. The real-world evidence we gained revealed metformin’s potential in ameliorating PD progression. In conclusion, our findings illuminated distinct PD pace subtypes with differing progression patterns, uncovered potential biological underpinnings driving different subtypes, and predicted repurposable drug candidates. This work may help better understand clinical and pathophysiological complexity of PD progression and accelerate precision medicine.

## Introduction

Parkinson’s disease (PD) is a progressive neurodegenerative disorder characterized by changes in both motor and non-motor functions and involves degeneration of multiple basal ganglia and cortical related circuits. PD is the second most prevalent neurodegenerative disorder, impacting approximately 2-3% of individuals aged over 65.^1,2^ The prevalence of the disease increases with advancing age, and it has emerged as a prominent health concern for the aging population.^1,2^ PD is characterized by the loss of dopamine-producing (dopaminergic) neurons in the substantia nigra and the accumulation of *α* - synuclein aggregates across multiple brain circuits and regions^1,2^. However, the precise etiological and pathological mechanisms underlying PD remain elusive. Consequently, as of now, there are no approved disease-modifying treatments known to slow, prevent, or reverse the progression of PD^3^.

In the past decades, there has been increasing recognition that “Parkinson’s disease” is not a single entity, but rather multiple sub-disorders classified under the “Parkinson’s disease” term with multiple overlapping etiologies^4–6^, leading to distinct progression trajectories during the PD course. Past clinical trials have struggled to account for the considerable heterogeneity in symptoms and progression and the pathophysiology underlying this clinical heterogeneity.^7^ This clinical and progression heterogeneity may have contributed to PD clinical trial failures.^7,8^ Administering the same medication to patients who share a PD clinical diagnosis but possess diverse underlying pathobiological processes, is unlikely to ameliorate disease progression. Moreover, without proper characterization of phenotypic variations, evaluating treatment response in clinical trials for PD is challenging. In this context, segmenting the heterogeneous population of a disease into relatively pathologically and biologically homogeneous subgroups, i.e., so-called subtypes, has shown significant promise for precision medicine and drug development. Recently, PD research has begun to more seriously shift its focus towards identifying PD subtypes.^8–11^ Previous studies have utilized various approaches for subtyping PD, such as determining subtypes based on PD risk genotypes, categorizing subtypes based on motor (e.g., postural instability and gait difficulty [PIGD]) or non-motor manifestations (e.g., mild cognitive decline subtype), or by employing emerging data-driven methods.^10,11^ However, despite the progress made, there is yet no consensus on a single subtyping method that effectively aids in PD therapy^10^ and accounts for disease progression^12–14^. One potential reason has been the insufficient appreciation and consideration given to PD progression heterogeneity. Approaches that utilize longitudinal data, beginning from a specific disease duration or disease milestone (e.g., early stage), could offer new insights in addressing PD heterogeneity.^15^

Increasing efforts have been dedicated to exploring the complex pathological and biological underpinnings of PD.^1,2,16,17^ These efforts have uncovered crucial mechanisms involved in PD, such as the aggregation of *α*-synuclein, neuroinflammation, mitochondrial dysfunction, and oxidative stress.^16–18^ Additionally, associations have been established between specific genes and the disease manifestations although notably there has been substantial heterogeneity even in monogenetic causes of PD.^19–21^ Most data have focused predominantly on the disease state of PD, offering limited insight into the disease’s progression. A few recent genetic studies have identified risk loci associated with motor and non-motor progression in PD.^22,23^ However, our understanding of the pathophysiological mechanisms that drive the heterogeneous progression of PD remains incomplete. This knowledge gap is an obstacle to discovering effective disease-modifying treatments that can slow, halt, or reverse PD progression. Further studies aimed at better understanding the complex interplay of factors influencing PD progression are needed to achieve the goal of developing efficacious therapeutic strategies.

In this study, our goal was to disentangle the clinical and progression heterogeneity of PD to accelerate precision medicine. To achieve this, we established an integrated data-driven framework that combines machine learning, deep learning, network medicine, and statistical approaches, enabling a multifaceted analysis of diverse data types (see Fig. 1). These included individual-level clinical records, biospecimens, neuroimaging, genetic and transcriptomic information, publicly available protein-protein interactome (PPI) and transcriptomics-based drug-gene signature data^24^, as well as patient-level real-world data (RWD). First, we characterized individual’s high-dimensional phenotypic progression data to uncover PD subtypes. This led to the identification of three pace subtypes of PD that exhibited distinct phenotypic progression patterns and were stable over time. Next, we identified indicative cerebrospinal fluid (CSF) and neuroimaging markers of the subtypes we discovered. Furthermore, by interrogating genetic and transcriptomic data with network medicine approaches, we identified subtype-specific molecular modules, revealing potential pathophysiological underpinnings driving the subtypes with distinct progression trends. Finally, we predicted potential therapeutic candidates by targeting subtype-specific molecular modules and estimated treatment effects of the candidates using real-world evidence (RWE) via analysis of two large-scale RWD databases. Our data suggested metformin’s potential in mitigating PD progression, as 1) it could counteract molecular alterations triggered in the rapid pace subtype, and 2) it was associated with an improved PD progression based on RWE.

**Figure 1.**
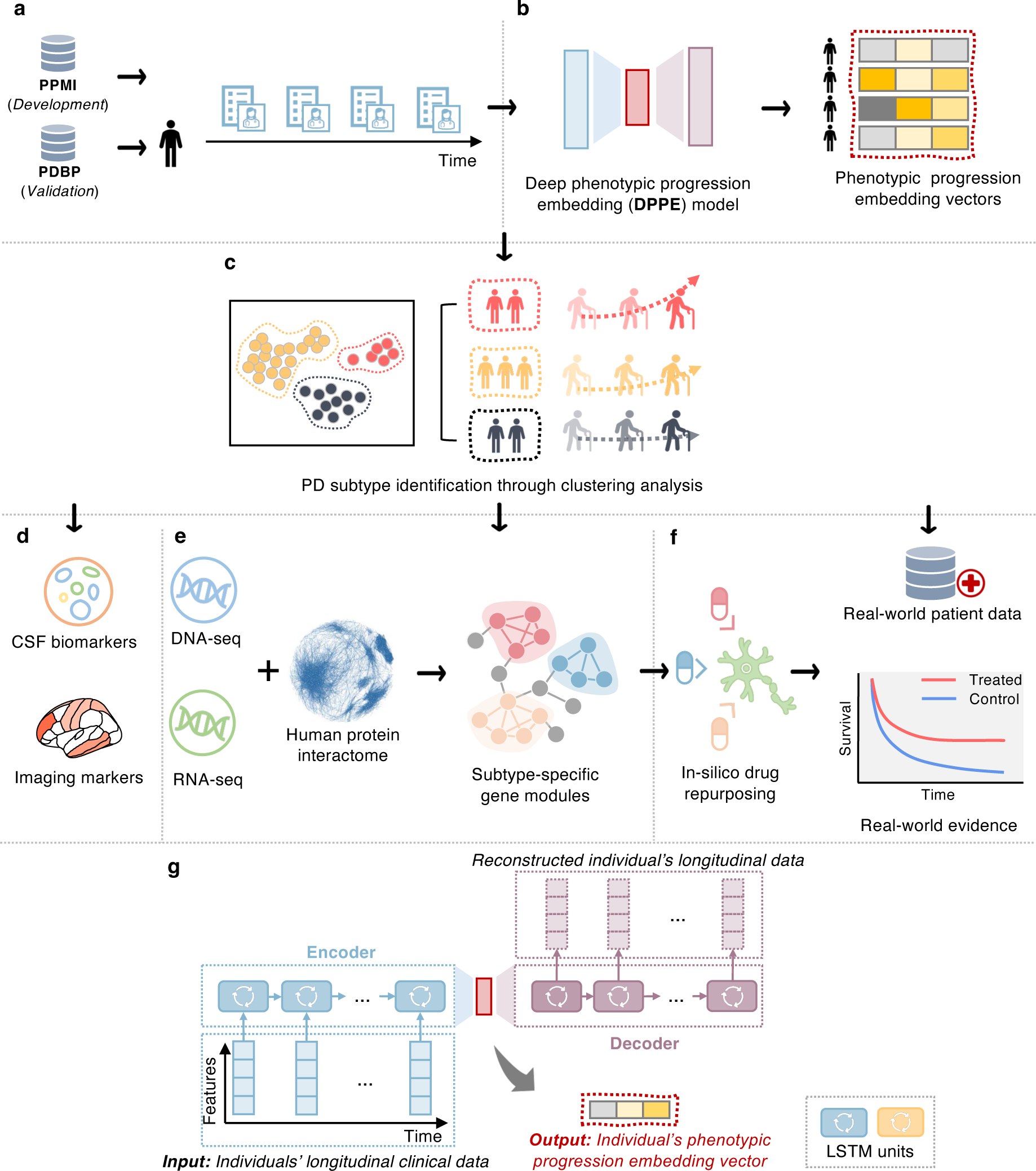
A diagram illustrating the present analysis. **a.** Collecting longitudinal clinical data from the PPMI and PDBP cohorts and conducting necessary data cleaning and preprocessing. **b.** Development of a deep phenotypic progression embedding (DPPE) model to learn a progression embedding vector for each individual, which encodes his/her PD symptom progression trajectory. **c.** Cluster analysis with the learned embedding vectors to identify PD subtypes, each of which reveal a unique PD progression pattern. **d.** Identifying CSF biomarkers and imaging markers the discovered PD subtypes. **e.** Construction of PD subtype-specific molecular modules based on genetic and transcriptomic data, along with human protein-protein interactome (PPI) network analyses, using network medicine approaches. **f.** In-silico drug repurposing based on subtype-specific molecular profiles and validation of drug candidates’ treatment efficiency based on analysis of large-scale real-world patient databases, i.e., the INSIGHT and OneFlorida+. **g.** Architecture of the DPPE model. Abbreviations: LSTM = the Long-Short Term Memory model; PD = Parkinson’s disease; PDBP = the Parkinson Disease Biomarkers Program; PPMI = the Parkinson’s Progression Markers Initiative.

## Results

### Study cohorts

In this investigation, we adopted data of participants in the PPMI study, an international observational PD study that systematically collected clinical, biospecimen, multi-omics, and brain imaging data of participants^25^. Our analysis included 406 *de novo* PD participants (PD diagnosis within the last 2 years and untreated at enrollment) in the PPMI cohort, comprising 140 (34.5%) women and 266 (65.5%) men, with an average age of 59.6±10.0 years at PD onset; 188 healthy control (HC) volunteers, comprising 67 (35.6%) women and 121 (64.4%) men; and 61 participants who had dopamine transporter scans without evidence of dopaminergic deficit (SWEDD), comprising 23 (37.7%) and 38 (62.3%) men (see Supplementary Table 1). Specifically, we trained deep learning model to capture PD phenotypic progression trends using 5 years longitudinal clinical assessments of *de novo* PD, HC, and SWEDD participants. Clustering analysis was conducted based on the learned progression profiles among the *de novo* PD participants to derive subtypes. We further examined individual’s neuroimaging, CSF, genetic, and transcriptomic data to identify subtype-specific biomarkers and molecular modules. To demonstrate robustness of our method in capturing PD progression trends to identify subtypes, we replicated our deep learning model among participants in the Parkinson Disease Biomarkers Program (PDBP)^26^. More details of participants included in this study can be found in the Methods and Supplementary Table 1.

### Discovery of three pace subtypes of de novo PD

We used individuals’ five years longitudinal records in over 140 items of diverse motor and non-motor assessments (see Supplementary Table 2). We built a deep learning model, termed deep phenotypic progression embedding (DPPE), for holistically modeling such multidimensional, longitudinal progression data of the participants (see Fig. 1g). The DPPE extended our previous work^27^, integrating a long short-term memory (LSTM)^8,13^ network with an autoencoder architecture. The LSTM is a specialized deep neural network designed for multivariate time sequence data analysis^28,29^. Specifically, DPPE had two components: 1) an encoder that used a LSTM to receive the longitudinal clinical data of each participant as input, capturing his/her phenotypic progression profile, and map it into a compact embedding vector; and 2) conversely, a decoder that employed another LSTM to unpack this compact embedding vector to reconstruct the individual’s raw input data. In this way, the DPPE was trained in an unsupervised manner by minimizing the difference between the input raw data and the reconstructed output. For training DPPE, we leveraged data of *de novo* PDs, HCs, and SWEDD individuals in the PPMI (see Supplementary Table 1). The trained model can generate a machine-readable embedding vector for each individual, encoding his/her phenotypic progression profile. More details of the DPPE model can be found in Fig. 1g and Methods section.

Next, we conducted cluster analysis based on the individuals’ DPPE-learned embedding vectors to identify PD subtypes. We used the agglomerative hierarchical clustering (AHC) algorithm with Euclidean distance calculation and Ward linkage criterion, because has demonstrated to be robust to different types of data distributions^30^. An issue of clustering analysis is how to determine the optimal cluster number. To this end, we considered 1) cluster separation in the dendrogram produced by the AHC, 2) 18 cluster structure measurements using the ‘NbClust’ software^31^, 3) cluster separation in the 2-dimensional (2D) space based on the t-distributed stochastic neighbor embedding (t-SNE) algorithm^32^, and 4) clinical interpretations of clusters. Using our method, three distinct subtypes were identified (see Supplementary Fig. 1 and Supplementary Results). The subtypes’ demographics and clinical characteristics at baseline and follow-up were detailed in Table 1 and Supplementary Tables 3 and 4, respectively. Subtype-specific averaged symptom progression trajectories and annual progression rates in terms of each single clinical assessment – as estimated by linear mixed effect models (adjusting for age, sex, and levodopa equivalent dose (LED) usage at visit) – were illustrated in Fig. 2a and summarized in Table 2, respectively. The specific characteristics of each subtype were elaborated upon below.

**Figure 2.**
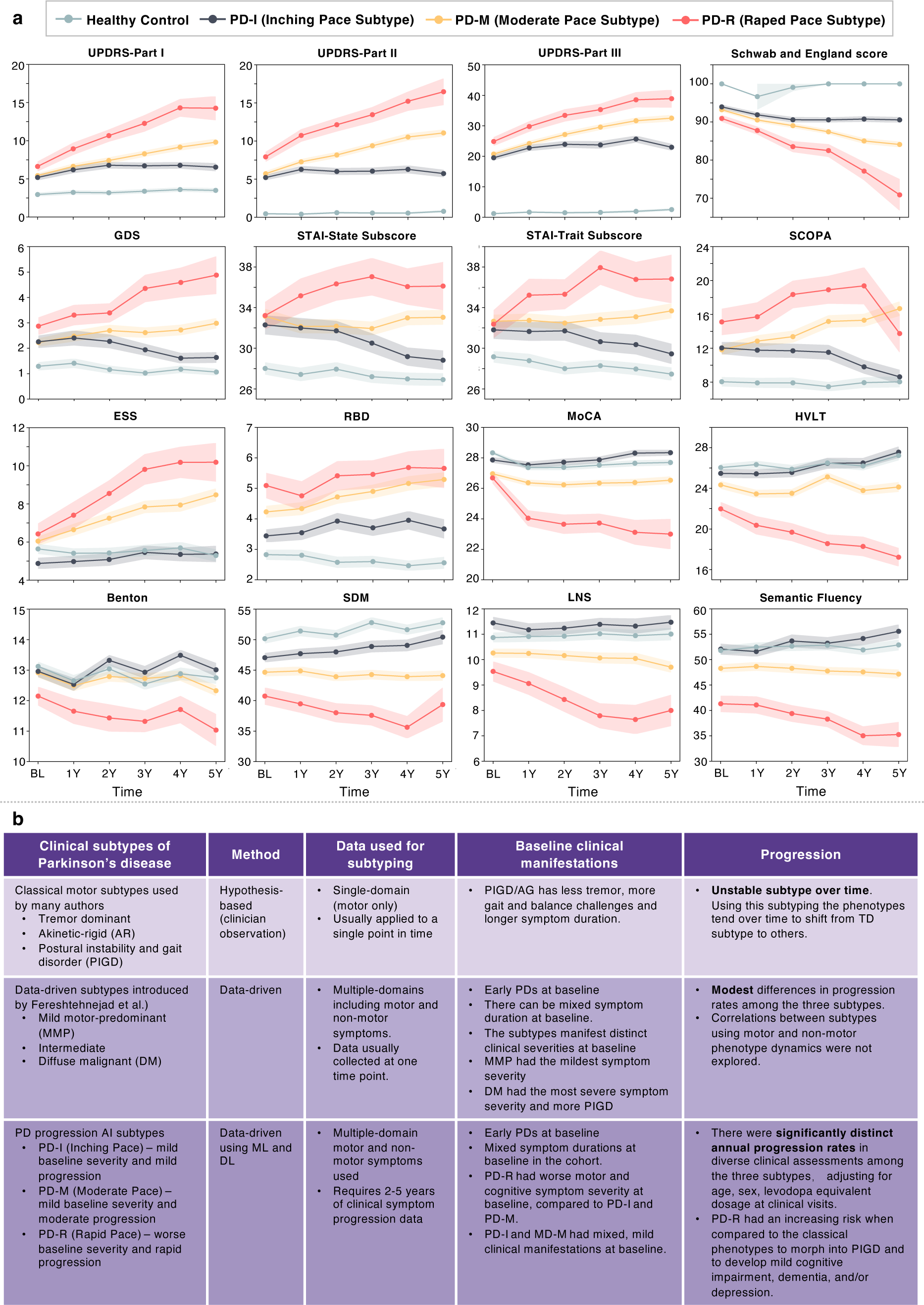
Progression patterns of the three PD subtypes. **a.** Averaged progression trajectories in clinical manifestations by subtypes. **b.** Comparisons of the identified pace subtypes with conventional motor subtypes and prior data-driven subtypes. Abbreviations: ESS = Epworth sleepiness score; GDS = Geriatric depression scale; HVLT = Hopkins Verbal Learning Test; LNS = Letter-number sequencing; MCI = mild cognition impairment; MDS-UPDRS = Movement Disorders Society–revised Unified Parkinson’s Disease Rating Scale; MoCA = Montreal Cognitive Assessment; PIGD = postural instability and gait disorder; PPMI = the Parkinson’s Progression Markers Initiative; RBD = REM sleep behavior disorder; SCOPA = Scales for Outcomes in Parkinson’s Disease; SDM = Symbol–Digit Matching; STAI = State-Trait Anxiety Inventory.

**Table 1.** Demographics and baseline clinical characteristics by subtypes in the PPMI cohort.

| Variables | Subtype PD-I<br>(Inching Pace) | Subtype PD-M<br>(Moderate Pace) | Subtype PD-R<br>(Rapid Pace) | P-value<sup>a</sup> | Post-hoc<sup>b</sup> | P-value<br>adjusted<sup>c</sup> |
| --- | --- | --- | --- | --- | --- | --- |
| **# of participants** | **145** | **207** | **54** | - | - | - |
| Age at onset, year, mean (SD) | 58.1 (9.9) | 59.4 (10.0) | 64.4 (8.6) | <0.001 | III vs. rest | - |
| Sex male, N (%) | 67 (46.2) | 155 (74.9) | 44 (81.5) | <0.001 | - | - |
| Race white, N (%) | 140 (96.6) | 196 (94.7) | 50 (92.6) | 0.422 | - | - |
| Symptom duration, month, mean (SD) | 0.593 (0.682) | 0.546 (0.742) | 0.667 (0.752) | 0.526 | - | - |
| Genetic risk score<sup>d</sup>, mean (SD) | -0.009 (0.004) | -0.009 (0.004) | -0.01 (0.004) | 0.615 | - | 0.879 |
| Family history, N (%) | 140 (96.6) | 196 (94.7) | 50 (92.6) | 0.711 | - | - |
| Education history group, N (%) |  |  |  |  | - | - |
| < 12 years | 10(6.9) | 9(4.4) | 7(13.0) | 0.102 | - | - |
| 12-16 years | 90(62.1) | 132(63.8) | 26(48.2) |  |  |  |
| > 16 years | 45(31.0) | 66(31.9) | 21(38.9) |  |  |  |
| ***Motor manifestations*** |  |  |  |  |  |  |
| MDS-UPDRS Part II, mean (SD) | 5.2 (3.7) | 5.7 (4.2) | 7.9 (4.4) | <0.001 | III vs. rest | <0.001 |
| MDS-UPDRS Part III, mean (SD) | 19.5 (8.8) | 20.7 (8.4) | 24.8 (9.6) | <0.001 | III vs. rest | 0.005 |
| H&Y Stage, mean (SD) | 1.55 (0.51) | 1.54 (0.49) | 1.70 (0.49) | 0.093 | - | 0.378 |
| Schwab and England score, mean (SD) | 93.9 (5.9) | 93.2 (5.6) | 90.9 (5.7) | 0.005 | III vs. rest | 0.004 |
| Tremor score, mean (SD) | 0.46 (0.32) | 0.50 (0.31) | 0.55 (0.34) | 0.14 | - | 0.252 |
| PIGD score, mean (SD) | 0.22 (0.25) | 0.20 (0.19) | 0.34 (0.26) | <0.001 | III vs. rest | <0.001 |
| Motor phenotype, N (%) |  |  |  |  |  |  |
| Tremor | 98 (67.6) | 154 (74.4) | 33 (61.1) | 0.235 | - | - |
| Indeterminate | 18 (12.4) | 22 (10.6) | 6 (11.1) |  |  |  |
| PIGD | 29 (20.0) | 31 (15.0) | 15 (27.8) |  |  |  |
| ***Non-motor manifestations*** |  |  |  |  |  |  |
| MDS-UPDRS Part I, mean (SD) | 5.2 (4.2) | 5.4 (3.8) | 6.6 (4.4) | 0.076 | - | 0.034 |
| Hallucination, mean (SD) | 0.01 (0.12) | 0.03 (0.18) | 0.04 (0.19) | 0.467 | - | 0.304 |
| Apathy, mean (SD) | 0.18 (0.51) | 0.21 (0.48) | 0.20 (0.49) | 0.82 | - | 0.657 |
| Pain, mean (SD) | 0.67 (0.83) | 0.72 (0.83) | 0.65 (0.84) | 0.806 | - | 0.426 |
| Fatigue, mean (SD) | 0.56 (0.71) | 0.67 (0.79) | 0.74 (0.84) | 0.272 | - | 0.058 |
| Sleep, mean (SD) |  |  |  |  |  |  |
| Epworth sleepiness score | 4.9 (3.4) | 6.0 (3.1) | 6.4 (3.8) | 0.001 | I vs. rest | 0.002 |
| REM sleep behavior disorder score | 3.4 (2.4) | 4.2 (2.7) | 5.1 (2.9) | <0.001 | I vs. rest | <0.001 |
| Sleep phenotype, missing = 1, N (%) |  |  |  |  |  |  |
| REM sleep behavior disorder positive | 38 (26.2) | 84 (40.8) | 26 (48.2) | 0.003 | - | - |
| REM sleep behavior disorder negative | 107 (73.4) | 122 (59.2) | 28 (51.8) |  |  |  |
| QUIP (Impulse control disorders) | 0.27 (0.62) | 0.31 (0.64) | 0.17 (0.46) | 0.284 | - | 0.360 |
| Geriatric depression scale | 2.26 (2.79) | 2.23 (2.801) | 2.87 (2.41) | 0.212 | - | 0.078 |
| Depression phenotype, missing = 1, N (%) |  |  |  |  |  |  |
| Normal | 125 (86.2) | 181 (87.9) | 42 (77.8) | 0.295 | - | - |
| Mild | 9 (6.2) | 17 (8.3) | 8 (14.8) |  |  |  |
| Moderate | 9 (6.2) | 7 (3.4) | 4 (7.4) |  |  |  |
| Severe | 2 (1.4) | 1 (0.5) | 0 (0) |  |  |  |
| State trait anxiety index, mean (SD) |  |  |  |  |  |  |
| State subscore | 32.3 (10.8) | 33.2 (9.9) | 33.2 (9.5) | 0.716 | - | 0.318 |
| Trait subscore | 31.8 (10.3) | 32.7 (8.8) | 32.4 (9.7) | 0.713 | - | 0.107 |
| SCOPA autonomic questionnaire, mean (SD) |  |  |  |  |  |  |
| Gastrointestinal (up+down) | 1.81 (1.93) | 2.1 (1.95) | 3.13 (2.35) | <0.001 | III vs. rest | <0.001 |
| Urinary | 3.88 (2.63) | 4.21 (2.89) | 5.41 (7.45) | 0.040 | I vs. III | 0.198 |
| Cardiovascular | 0.39 (0.68) | 0.51 (0.86) | 0.52 (0.66) | 0.309 | - | 0.316 |
| Thermoregulatory | 0.46 (0.87) | 0.47 (0.84) | 0.24 (0.72) | 0.19 | - | 0.340 |
| Pupillomotor | 0.35 (0.58) | 0.42 (0.64) | 0.50 (0.79) | 0.289 | - | 0.182 |
| Skin | 0.69 (0.88) | 0.68 (0.91) | 0.83 (0.96) | 0.53 | - | 0.243 |
| Sexual | 4.47 (6.48) | 3.43 (5.68) | 4.48 (6.71) | 0.233 | - | 0.629 |
| Total (sum all) | 12.04 (8.39) | 11.81 (8.32) | 15.11 (11.16) | 0.044 | II vs. III | 0.020 |
| Cognitive function, mean (SD) |  |  |  |  |  |  |
| MoCA-visuospatial | 4.6 (0.8) | 4.5 (0.8) | 4.4 (0.8) | 0.23 | - | 0.271 |
| MoCA-naming | 3.0 (0.2) | 2.9 (0.3) | 2.9 (0.4) | 0.043 | I vs. III | 0.135 |
| MoCA-attention | 5.8 (0.5) | 5.8 (0.6) | 5.7 (0.7) | 0.532 | - | 0.393 |
| MoCA-language | 2.7 (0.6) | 2.5 (0.7) | 2.6 (0.7) | 0.013 | I vs. II | 0.011 |
| MoCA-delayed recall | 3.7 (1.3) | 3.3 (1.4) | 2.9 (1.6) | <0.001 | I vs. rest | 0.043 |
| MoCA total score | 27.9 (2.0) | 27.0 (2.4) | 26.7 (2.4) | <0.001 | I vs. rest | 0.004 |
| Benton judgment of line orientation | 13.0 (2.1) | 12.9 (2.1) | 12.1 (2.2) | 0.036 | III vs. rest | 0.020 |
| HVLT-total recall | 25.5 (4.6) | 24.3 (5.1) | 22.0 (4.7) | <0.001 | III vs. rest | 0.023 |
| HVLT-delayed recall | 8.8 (2.4) | 8.3 (2.5) | 7.2 (2.5) | <0.001 | III vs. rest | 0.016 |
| HVLT-discrimination recognition | 10.1 (2.3) | 9.7 (2.5) | 8.5 (2.9) | <0.001 | III vs. rest | 0.572 |
| HVLT-retention | 0.9 (0.2) | 0.8 (0.2) | 0.8 (0.2) | 0.072 | - | 0.073 |
| LNS | 11.4 (2.7) | 10.3 (2.4) | 9.5 (2.8) | <0.001 | I vs. rest | <0.001 |
| Semantic fluency | 52.1 (12.1) | 48.3 (10.5) | 41.3 (11.4) | <0.001 | All comparisons | <0.001 |
| Symbol digit test | 47.0 (8.6) | 44.6 (9.0) | 40.7 (9.9) | <0.001 | All comparisons | <0.001 |
| Cognitive phenotype, missing = 7, N (%) |  |  |  |  |  |  |
| Normal | 135 (97.8) | 200 (96.6) | 41 (75.9) | <0.001 | - | - |
| MCI | 2 (1.4) | 4 (1.9) | 8 (14.8) |  |  |  |
| Dementia | 1 (0.7) | 3 (1.5) | 5 (9.3) |  |  |  |
| <sup>a</sup> P-values were calculated using ANOVA (for continuous variables) and $\chi^2$ test (for categorical variables) where appropriate.<br><sup>b</sup> Post-hoc analysis was performed using the Tukey HSD test when the ANOVA P-value < 0.05.<br><sup>c</sup> ANCOVA was used to calculate p-values (for continuous variables) adjusting for age, sex, and levodopa equivalent daily dose.<br><sup>d</sup> Genetic risk score were calculated based on 90 PD-related loci reported in the latest Genome wide association study<sup>33</sup>.<br>Abbreviations: HVLT = Hopkins Verbal Learning Test; LNS = Letter-number sequencing; MCI = mild cognitive impairment; MDS-UPDRS = Movement Disorders Society–revised Unified Parkinson’s Disease Rating Scale; MoCA = Montreal Cognitive Assessment; PIGD = postural instability and gait disorder; PPMI = the Parkinson’s Progression Markers Initiative; SCOPA = Scales for Outcomes in Parkinson’s Disease. | | | | | | |

**Table 2.** Annual progression rates in clinical manifestations and CSF biomarkers by subtypes assessed by linear mixed effects models in the PPMI cohort.

| Variable | Subtype PD-I<br>(Inching Pace) |  | Subtype PD-M<br>(Moderate Pace) |  | Subtype PD-R<br>(Rapid Pace) |  |
| --- | --- | --- | --- | --- | --- | --- |
| | $\beta$ | *P* value | $\beta$ | *P* value | $\beta$ | *P* value |
| ***Motor manifestations*** |  |  |  |  |  |  |
| UPDRS-Part II | 0.42 (0.24, 0.60) | <0.001 | 1.02 (0.88, 1.16) | <0.001 | 1.98 (1.52, 2.44) | <0.001 |
| UPDRS-Part III | 1.03 (0.62, 1.43) | <0.001 | 2.07 (1.74, 2.40) | <0.001 | 3.08 (2.12, 4.03) | <0.001 |
| H&Y Stage | 0.07 (0.05, 0.09) | <0.001 | 0.09 (0.08, 0.10) | <0.001 | 0.15 (0.10, 0.20) | <0.001 |
| Schwab and England score | -0.98 (-1.26, -0.69) | <0.001 | -1.73 (-1.98, -1.48) | <0.001 | -3.89 (-4.93, -2.85) | <0.001 |
| Tremor score | 0 (-0.02, 0.01) | 0.917 | 0.03 (0.02, 0.04) | <0.001 | 0 (-0.03, 0.02) | 0.842 |
| PIGD score | 0.04 (0.02, 0.05) | <0.001 | 0.06 (0.04, 0.07) | <0.001 | 0.19 (0.13, 0.24) | <0.001 |
| ***Non-motor manifestations*** |  |  |  |  |  |  |
| UPDRS-Part I | 0.54 (0.45, 0.62) | <0.001 | 0.88 (0.80, 0.96) | <0.001 | 1.70 (1.53, 1.88) | <0.001 |
| Hallucination | 0.02 (0.01, 0.03) | <0.001 | 0.03 (0.02, 0.04) | <0.001 | 0.11 (0.06, 0.16) | <0.001 |
| Apathy | 0.01 (-0.01, 0.03) | 0.219 | 0.05 (0.03, 0.07) | <0.001 | 0.12 (0.06, 0.18) | <0.001 |
| Pain | 0.03 (0, 0.06) | 0.026 | 0.06 (0.04, 0.09) | <0.001 | 0.14 (0.10, 0.18) | <0.001 |
| Fatigue | 0.05 (0.02, 0.07) | <0.001 | 0.08 (0.06, 0.11) | <0.001 | 0.18 (0.13, 0.23) | <0.001 |
| Sleep |  |  |  |  |  |  |
| Epworth sleepiness score | 0.17 (0.05, 0.30) | 0.008 | 0.49 (0.37, 0.60) | <0.001 | 1.07 (0.79, 1.35) | <0.001 |
| REM sleep behavior disorder | 0.18 (0.09, 0.28) | <0.001 | 0.27 (0.19, 0.34) | <0.001 | 0.14 (-0.01, 0.32) | 0.076 |
| QUIP (Impulse control disorders) | 0.02 (-0.01, 0.04) | 0.232 | 0.06 (0.03, 0.08) | <0.001 | 0.02 (-0.02, 0.06) | 0.332 |
| Geriatric depression scale | -0.12 (-0.21, -0.02) | 0.014 | 0.11 (0.04, 0.19) | 0.002 | 0.47 (0.25, 0.69) | <0.001 |
| State trait anxiety index |  |  |  |  |  |  |
| State subscore | -0.53 (-0.83, -0.24) | <0.001 | -0.02 (-0.27, 0.22) | 0.851 | 0.81 (0.24, 1.40) | 0.008 |
| Trait subscore | -0.32 (-0.58, -0.02) | 0.020 | 0.16 (-0.07, 0.39) | 0.172 | 1.09 (0.50, 1.69) | <0.001 |
| SCOPA autonomic questionnaire |  |  |  |  |  |  |
| Gastrointestinal | 0.28 (0.19, 0.36) | <0.001 | 0.31 (0.25, 0.37) | <0.001 | 0.39 (0.25, 0.54) | <0.001 |
| Urinary | 0.12 (0.04, 0.20) | 0.004 | 0.28 (0.17, 0.38) | <0.001 | 0.34 (-0.07, 0.76) | 0.111 |
| Cardiovascular | 0.05 (0.02, 0.09) | 0.002 | 0.07 (0.04, 0.09) | <0.001 | 0.21 (0.13, 0.29) | <0.001 |
| Thermoregulatory | 0.02 (-0.02, 0.06) | 0.254 | 0.06 (0.03, 0.09) | <0.001 | 0.07 (0.02, 0.12) | 0.017 |
| Pupillomotor | 0.02 (0, 0.05) | 0.057 | 0.04 (0.02, 0.06) | <0.001 | 0.07 (0.02, 0.13) | 0.006 |
| Skin | 0.03 (0.00, 0.07) | 0.051 | 0.10 (0.07, 0.13) | <0.001 | 0.14 (0.06, 0.23) | 0.002 |
| Sexual | 0.14 (-0.08, 0.37) | 0.216 | 0.37 (0.21, 0.53) | <0.001 | 0.69 (0.30, 1.07) | 0.001 |
| Total | 0.68 (0.42, 0.94) | <0.001 | 1.23 (0.97, 1.47) | <0.001 | 1.88 (1.15, 2.61) | <0.001 |
| **Cognitive function** |  |  |  |  |  |  |
| MoCA | 0.05 (-0.02, 0.13) | 0.170 | -0.09 (-0.17, -0.02) | 0.018 | -0.99 (-1.32, -0.67) | <0.001 |
| Benton judgment of line orientation | 0.02 (-0.04, 0.08) | 0.548 | -0.06 (-0.12, -0.01) | 0.026 | -0.27 (-0.41, -0.13) | <0.001 |
| HVLT-total recall | 0.22 (0.05, 0.38) | 0.006 | 0.03 (-0.10, 0.17) | 0.674 | -0.98 (-1.24, -0.72) | <0.001 |
| HVLT-delayed recall | 0.11 (0.04, 0.19) | 0.005 | 0.01 (-0.06, 0.08) | 0.735 | -0.55 (-0.74, -0.34) | <0.001 |
| HVLT-discrimination recognition | 0.23 (0.14, 0.32) | <0.001 | 0.16 (0.09, 0.23) | <0.001 | 0.05 (-0.14, 0.25) | 0.602 |
| HVLT-retention | 0 (0, 0.01) | 0.206 | 0 (0, 0) | 0.575 | -0.05 (-0.07, -0.03) | <0.001 |
| LNS | -0.10 (-0.17, -0.02) | 0.017 | -0.10 (-0.17, -0.04) | 0.002 | -0.48 (-0.68, -0.28) | <0.001 |
| Semantic fluency | 0.25 (-0.11, 0.61) | 0.183 | -0.23 (-0.47, 0.01) | 0.058 | -1.46 (-2.07, -0.84) | <0.001 |
| Symbol digit test | 0.34 (-0.04, 0.71) | 0.081 | -0.12 (-0.37, 0.12) | 0.335 | -1.23 (-1.90, -0.56) | <0.001 |
| For each variable, we fitted a linear mixed effect model for each PD subtype, specifying time (year) as the explanatory variable of interest. For all models, individual variation was included as a random effect, and age, sex, and levodopa equivalent dose were included as covariates. The annual progression rate of a clinical assessment was obtained as the estimated as $\beta$ (95% CI) of time.<br>Abbreviations: HVLT = Hopkins Verbal Learning Test; LNS = Letter-number sequencing; MDS-UPDRS = Movement Disorders Society–Unified Parkinson’s Disease Rating Scale; MoCA = Montreal Cognitive Assessment; PIGD = postural instability and gait disorder; PPMI = the Parkinson’s Progression Markers Initiative; SCOPA = Scales for Outcomes in Parkinson’s Disease. |  |  |  |  |  |  |

**The Rapid Pace subtype (PD-R), marked by rapid symptom progression,** consisted of 54 (13.3%) individuals (see Fig. 2 and Tables 1 and 2). Compared to other subtypes, PD-R had more males (N=44 [81.5%]) with the highest average age at PD onset (64.4±8.6 years). Individuals of PD-R experienced more severe motor symptoms at baseline, as indicated by MDS-UPDRS Parts II and III and the Schwab and England scale, as well as more non-motor problems, especially cognitive impairment (see Table 2). Remarkably, PD-R was associated with the most rapid annual progression rates in most motor and non-motor symptoms among the three subtypes. For instance, compared to other subtypes, PD-R exhibited greater annual progression rates in motor assessments including MDS-UPDRS Part II (1.98/year, 95% CI [1.52, 2.44], *P* < 0.001) and Part III (3.08/year, 95% CI [2.12, 4.03], *P* < 0.001), and Schwab and England score (−3.89/year, 95% CI [−4.93, −2.85], *P* < 0.001). PD-R also showed the greatest annual increase rate regarding the overall non-motor function, measured by MDS-UPDRS Part I (1.70/year, 95% CI [1.53, 1.88], *P* < 0.001). Rapid progression rates were also observed in specific non-motor functions in PD-R such as sleep (e.g., Epworth sleepiness score and REM sleep behavior disorder), mood (e.g., State trait anxiety index and Geriatric depression scale), autonomic problem (Scales for Outcomes in Parkinson’s Disease autonomic questionnaire), and cognitive performance (e.g., MoCA) (see Fig. 2a and Table 2).

**The Inching Pace subtype (PD-I), characterized by mild baseline symptoms and mild symptom progression**, encompassed 145 participants (35.7%) (see Fig. 2 and Tables 1 and 2). Compared to others, PD-I had a relatively lower proportion of men (46.2%, N=67) and a younger age at PD onset (58.1±9.9 years). In addition, individuals of PD-I exhibited milder motor and non-motor symptoms at baseline. This was substantiated by their lower scores on the Movement Disorders Society – Unified Parkinson’s Disease Rating Scale (MDS-UPDRS) Parts II and III, and their less severe sleep and cognitive impairments (see Table 1). Furthermore, PD-I demonstrated the most gradual PD progression among all subtypes, indicated by the lowest annual progression rates in most PD symptoms estimated by the linear mixed effect models (see Fig. 2 and Table 2). Specifically, the progression rates of overall motor symptoms were low in MDS-UPDRS Part II (0.42/year, 95% CI [0.24, 0.60], *P* < 0.001) and Part III (1.03/year, 95% CI [0.62, 1.43], *P* < 0.001), and Schwab and England score (−0.98 /year, 95% CI [−1.26, −0.69], *P* < 0.001). Non-motor symptoms also progressed at a moderate rate, for example, MDS-UPDRS Part I of PD-I increased at a low rate of 0.54 (95% CI [0.45, 0.62], *P* < 0.001).

**The Moderate Pace subtype (PD-M), characterized by mild baseline symptoms and moderate symptom progression**, included 207 (50.9%) individuals (see Fig. 2 and Tables 1 and 2). This subtype had a higher proportion of men (N=155 [74.9%]) compared to PD-I, and these individuals presented with an average age of 59.4±10.0 years at PD onset. Although individuals of PD-M exhibited mild motor and non-motor symptoms at enrollment, mixed with PD-I, they demonstrated worse clinical symptoms since the second year of follow-up when compared to PD-I (see Supplementary Tables 3 and 4). Notably, compared to the PD-I, PD-M displayed faster (generally moderate level) progression rates in most clinical manifestations (see Fig. 2a and Table 2). In terms of overall motor symptoms, the MDS-UPDRS Part II score showed an annual increase of 1.02 (95% CI [0.88, 1.16], *P* < 0.001), the MDS-UPDRS Part III score had an annual increase of 2.07 (95% CI [1.74, 2.40], *P* < 0.001), and the Schwab and England score showed an annual change of −1.73 (95% CI [−1.98, −1.48], *P* < 0.001). For non-motor symptoms, for instance, MDS-UPDRS Part I exhibited an estimated annual increase of 0.88 (95% CI [0.80, 0.96], *P* < 0.001).

We further investigated phenotype alterations over time across our identified subtypes. Here we examined motor phenotypes (tremor dominant [TD], indeterminate, and PIGD) and non-motor phenotypes including cognition (normal cognition/mild cognitive impairment/dementia), REM sleep behavior disorder [RBD] (positive/negative), and depression (non/mild/moderate/severe depression) using Sankey diagrams (see Extended Data Fig. 1). As shown in Extended Data Fig. 1, the individuals of PD-R had an increasing risk of developing more advanced phenotypes. These included PIGD, cognitive dysfunction (mild cognition impairment [MCI] and dementia), and moderate-to-severe depression. Individual of PD-I and PD-M were likely to had stable phenotypes throughout the PD course.

#### The subtypes are reproducible in the PDBP cohort

Following the same procedure above based on clinical variables shared with PPMI, we also obtained the three-cluster structure using clinical progression data of early PD individuals in the PDBP cohort (see Supplementary Results). Clinical characteristics at baseline and follow-up, and PD symptom progression profiles of these re-identified subtypes closely mirrored those uncovered in our primary analysis using the PPMI cohort (see Extended Data Fig. 2 and Supplementary Tables 5-8). These demonstrated the reproducibility and robustness of our subtyping approach and validated the pace subtypes we identified in the PPMI cohort.

### Discovery of CSF biomarkers of the PD pace subtypes

We assessed cerebrospinal fluid (CSF) biomarkers among the identified subtypes at baseline and identified potential indicators of the pace subtypes (see Fig. 3a and Supplementary Table 9). While the phosphorylated tau (P-tau) and total tau (T-tau) levels differentiated each subtype from HCs, these biomarkers were not effective at differentiating among the subtypes. The P-tau/α-synuclein ratio might be the most efficacious biomarker for distinguishing among the three subtypes at baseline. Furthermore, the amyloid beta-42 (A*β*-42)/P-tau and A*β*-42/T-tau ratios showed potential in differentiating PD-R (rapid progression) from the other two subtypes. However, they faced challenges when attempting to distinguish PD-M against PD-I. This echoed the similarity in clinical manifestations of the two subtypes at baseline (see Fig. 2 and Tables 1 and 2).

**Figure 3.**
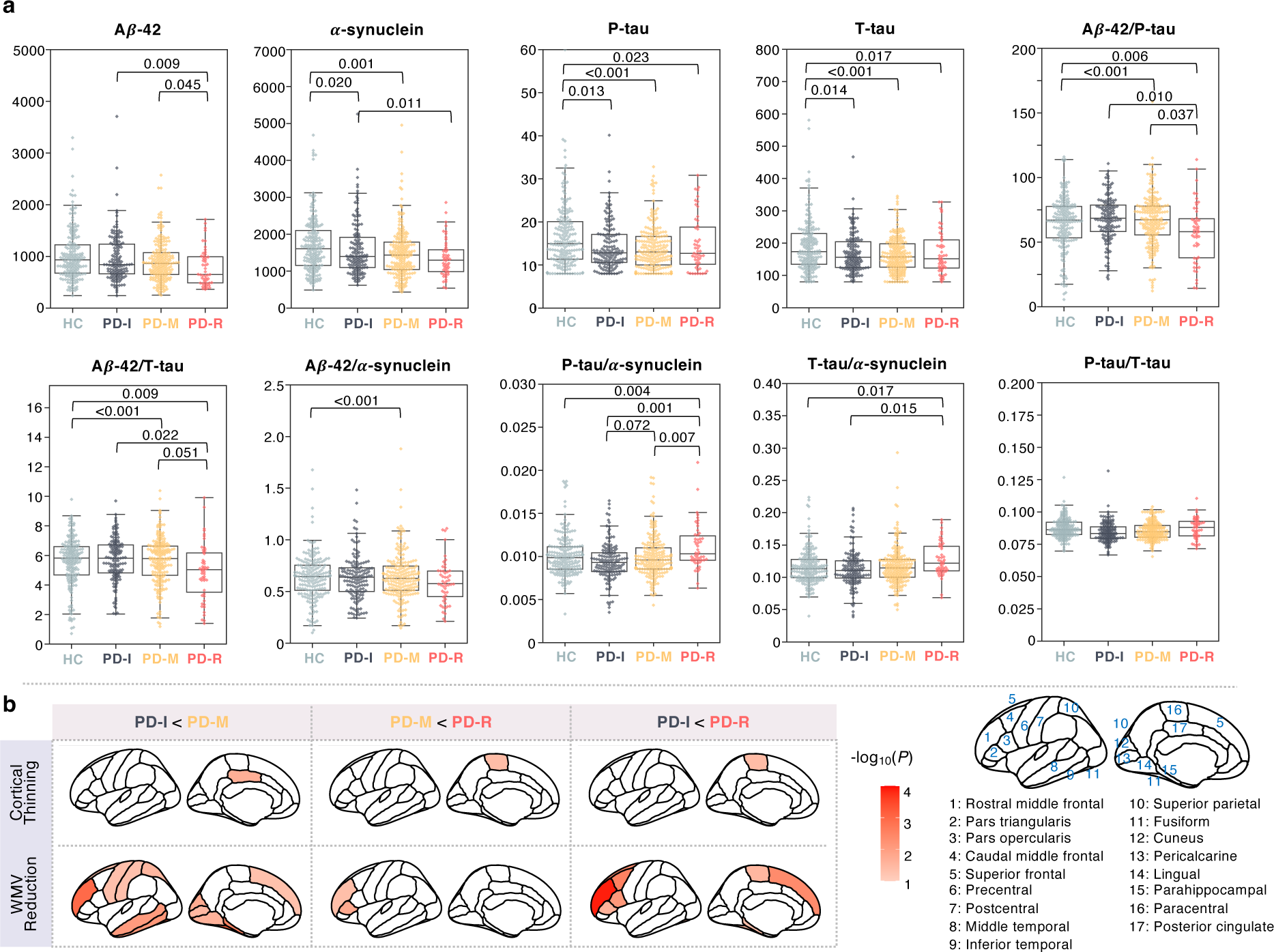
CSF biomarkers and neuroimaging markers of the identified subtypes. a. CSF biomarkers by PD subtypes. **b.** Regions showing significant signals in 1-year brain atrophy between a pair of subtypes. 1-year brain atrophy was measured by cortical thickness and white matter volume from 34 region of interests (ROIs), defined by the Desikan-Killiany atlas (averaged over the left and right hemispheres). Color density denotes significance in terms of −log_10_(*P*). Abbreviations: A*β*-42 = the 42 amino acid form of amyloid-*β*; CSF = cerebrospinal fluid.

### Discovery of imaging biomarkers of the PD pace subtypes

We examined brain atrophy measured by alterations of cortical thickness and white matter volume (WMV) across 34 brain regions of interest (ROIs) defined by the Desikan-Killiany atlas (averaged over the left and right hemispheres) within the first year after baseline. In Fig. 3b, we marked in red the ROIs where atrophy measures significantly differentiated each pair of PD subtypes (*P* < 0.05), with color intensity reflecting the significance level, i.e., −log_10_(*P*). We observed that reduction of WMV may be more useful than that of cortical thickness in differentiating the subtypes. Specifically, compared to PD-I, PD-M had significantly higher WVM reduction in certain ROIs, such as rostral middle frontal, superior frontal, precentral, postcentral, middle and inferior temporal, fusiform, lingual, parahippocampal gyri, superior parietal, cuneus, and pericalcarine, and cortical thickness atrophy in the posterior cingulate. Compared to PD-M, PD-R had more WVM reduction in rostral middle frontal gyrus and pars triangularis as well as cortical thickness reduction in paracentral gyrus. Compared to PD-I, PD-R had higher WVM reduction in multiple ROIs including rostral middle frontal, pars triangularis, pars opercularis, caudal middle frontal, superior frontal, paracentral, fusiform, and parahippocampal gyri, as well as cortical thickness of the paracentral gyrus. Together, these results indicated that increased WVM atrophy at earlier stage could serve as potential markers for the pace subtypes with distinct progression patterns.

### Discovery of genetic components of the PD pace subtypes

We conducted genetic data analyses to identify genetic factors associated to each PD subtype. Given the limited sample size, which makes a genome-wide association study (GWAS) unfeasible, we focused on 90 PD-related single-nucleotide polymorphisms (SNPs) reported in the latest GWAS study^33^ and apolipoprotein E (*APOE*) alleles, a known risk factor of AD. Similar to a previous study^34^, we utilized the hypergeometric tests to identify variants enriched in each subtype (see Methods). Our data suggested certain SNPs that could potentially be associated with each PD subtype (see Extended Data Fig. 3). Since impact of single genetic factors (i.e., SNPs) is low, we further conducted network-based analysis to amplify the genetic signals. We first mapped the subtype-associated SNPs to potential causal genes, leading to a list of genetic associated genes for each subtype. Next, in a human protein-protein interactome (PPI) network we built (see Methods), we located these genetic associated genes along with their connected PD contextual genes to construct a sub-network as the genetic molecular module of the subtype. The PD contextual genes were obtained through single nucleus RNA sequence (snRNA-seq) data from dopamine neurons. More details can be found in the Methods and Supplementary Methods.

We successfully identified genetic molecular module for each subtype (see Fig. 4a and Extended Data Fig. 4). For instance, 17 genes were identified as potential genetic associated genes of PD-R. Among these, 14 (*CNTNAP1*, *TRIM40*, *RETREG3*, *FYN*, *MBNL2*, *ATP6V0A1*, *STAT3*, *BECN1*, *BAG6*, *HLA-DRB1*, *HLA-DRB5*, *HLA-DQB1*, *SIPA1L2*, and *PRRC2A*) directly connected to snRNA-seq-derived PD contextual genes in the PPI network, thus constructing a genetic molecular module of PD-R, as depicted in Fig. 4a. Among the genes identified in the module was the signal transducer and activator of transcription 3 (*STAT3*), which transmits signals for the maturation of immune system cells. In microglia, *STAT3* is known to influence the expression of inflammatory genes.^35^ *FYN* is a member of the protein tyrosine kinase oncogene family, which plays a critical role in cell communication and signaling pathways, particularly in the nervous system. *FYN* can promote *STAT3* signaling as part of proinflammatory priming of microglia.^36^ *BECN1* (beclin 1) regulates autophagy, which clears toxic proteins such as *α*-synuclein aggregates in brain. *LRRK2* was found as a hub node in the modules of PD-I and PD-M. This was supported by previous evidence that mutated *LRRK2* is likely to be associated with lower progression rate of PD.^37^ Finally, pathway enrichment analyses suggested prominent pathways enriched in the three subtypes (see Fig. 4b and Extended Data Fig. 4b and d). For instance, some notable pathways were associated with the PD-R, including neuroinflammation (immune system-related pathways), oxidative stress, metabolism (insulin receptor signaling and Type I diabetes pathways), and the AD pathway. Top-ranked pathways for each subtype can be found in the Fig. 4b and Extended Data Fig. 4b and d and the full pathways lists can be found in the Extended Data File 1.

**Figure 4.**
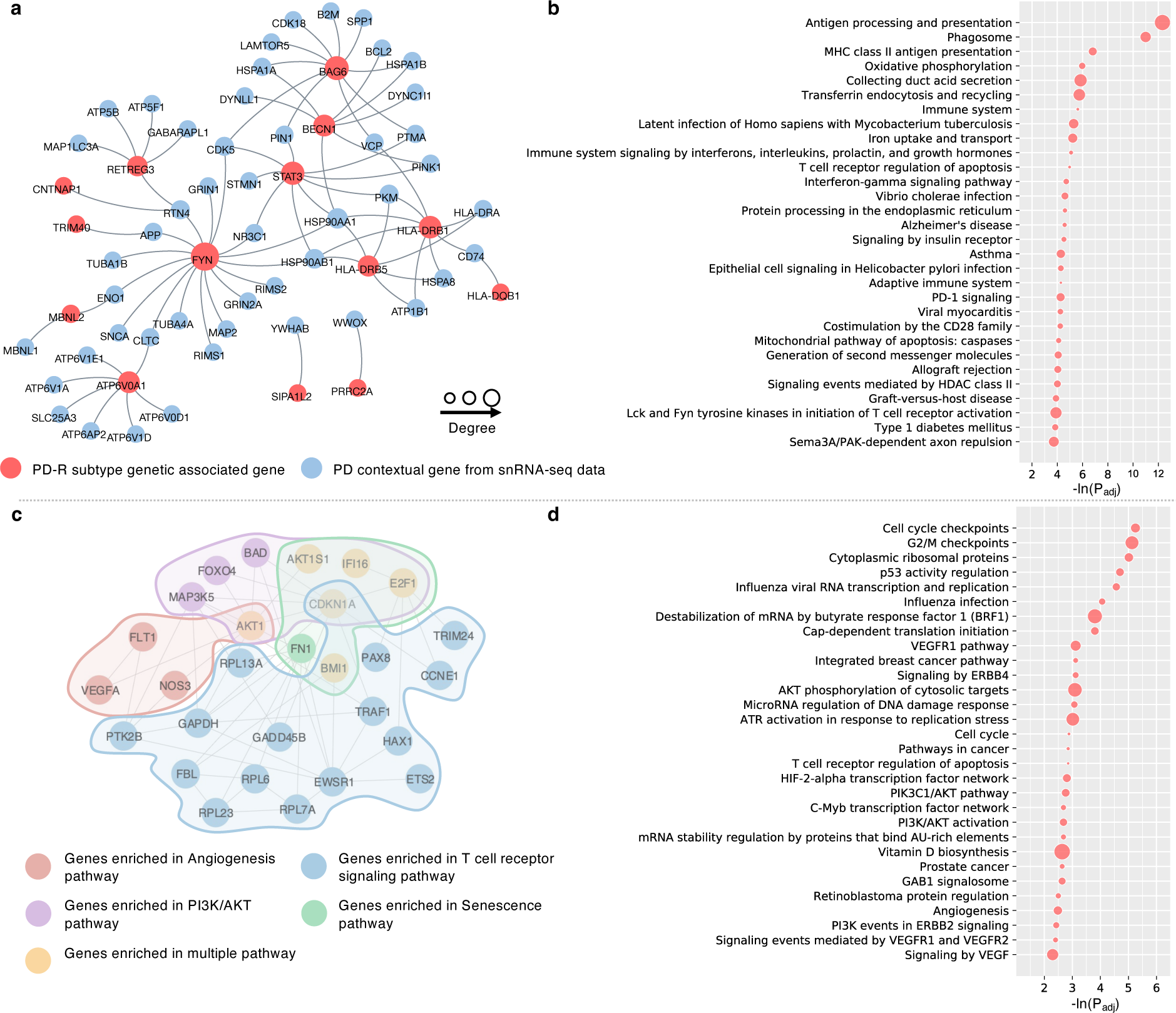
PD^R^ subtype-specific molecular modules revealing potential biological mechanisms of rapid PD progression. **a.** Genetic molecular module of PD-R. **b.** Pathways enriched based on genetic molecular module of PD-R. **c.** A sub-network of transcriptomic molecular module of PD-R. The entire transcriptomic molecular module of PD-R can be in the Extended Data File Fig. 8. **d.** Pathways enriched based on transcriptomic molecular module of PD-R.

### Discovery of transcriptomic profiles of the PD pace subtypes

We next investigated the transcriptomic changes associated with each PD subtype using the whole blood bulk RNA-seq data in the PPMI cohort (see Methods). Differential gene expression analyses, comparing each subtype with HCs, identified 2176, 2376, and 2305 differentially expressed genes (DEG, adjusted *P* < 0.05) for the PD-I, PD-M, and PD-R subtypes, respectively (see Extended Data Fig. 5 and Extended Data File 2).

Drawing on the DEGs of each subtype and PPI network we built as input, we utilized our Genome-wide Positioning Systems network (GPSnet) algorithm^38^ to identify transcriptomic molecular modules specific to each subtype (see Methods). Our analysis was grounded on the bases that a subtype-specific molecular module is a set of genes which are 1) densely interconnected within the PPI network and 2) differentially expressed in the specific subtype. We identified three distinct molecular modules for PD-I, PD-M, and PD-R, consisting of 211, 213, and 240 genes, respectively (see Extended Data Figs. 6-8). Focusing on PD-R, we found several genes of interest within the module (see Extended Data Fig. 8). For instance, the module gene *E2F1*, a member of the *E2F* family of transcription factors, has been previously linked to neuronal damage and death.^39,40^ The module gene apolipoprotein A1 (*APOA1*) produces the *APOA1* protein, a major protein component of high-density lipoprotein (HDL) in the blood. A previous PPMI cohort study^41^ has revealed the association between plasma *APOA1* level with age at PD onset and motor symptom severity. Another module gene, *NEDD4*, promotes removal of *α* - synuclein via a lysosomal process in human cells which may help resist PD.^42^ Furthermore, the *GATA* binding protein 2 (*GATA2*) has demonstrated its crucial role in neuronal development, specifically influencing the cell fate determination of catecholaminergic sympathetic neurons and modulating the expression of endogenous neuronal *α*-synuclein.^43^ Pathway enrichment analyses were conducted based on the subtype-specific modules and suggested pathways enriched in each subtype (see Figs. 4c and d and Extended Data Fig. 9). Taking PD-R as an example, Figure 4c illustrated a sub-network of its transcriptomic molecular module. The phosphoinositide-3-kinase/protein kinase B (PI3K/AKT), angiogenesis, T cell receptor signaling, and senescence pathways may play a role in rapid PD progression, i.e., PD-R. Top-ranked pathways in each subtype can be found in the Figs. 4c and d and Extended Data Fig. 9 and the full pathways lists were listed in Extended Data File 3.

### A classification model for distinguishing the PD pace subtypes at early stage

The identified PD pace subtypes demonstrated over 5-year progression trends. To gain more prognostic insights, we built a classification model for separating the pace subtypes at early stage. We used individuals’ data including demographics, genetic data, as well as clinical assessments, CSF biomarkers, and brain atrophy measures collected within the first year after baseline. The model was based on a cascade framework^44^ consisting of two base random forest classifiers: one separating PD-R from the others and another distinguishing PD-I and PD-M (see Extended Data Fig. 10). As shown in the figure, the model was very effective in separating PD-R from the other two subtypes, attaining an area under the receiver operating characteristics curve (AUC) of 0.87±0.05. While distinguishing PD-I and PD-M was challenging due to their similar clinical characteristics at baseline, the model still achieved acceptable performance with an AUC of 0.74±0.07.

### Discovery of PD pace subtype-based repurposable drug candidates

We sought to identify potential treatments to slow or halt PD progression by targeting the subtype-specific molecular bases. To achieve this goal, we used transcriptomics-based drug-gene signature data in human cell lines from the Connectivity Map (CMap) database^24^. We evaluated each drug’s potential therapeutic effects for preventing PD progression by assessing its ability to reverse dysregulated gene expression levels of the subtype-specific molecular modules (see Methods). More specifically, we used gene set enrichment analysis (GSEA) to compute an enrichment score (ES) with permutation tests for each tested drug.^45,46^ We chose ES > 3 and *Q* < 0.05 as the cutoffs to prioritize potential drug candidates (see Methods for more details). In total, we investigated 1,309 drugs from the CMap. Our analysis resulted in 49, 65, and 207 candidates (ES > 3 and *Q* < 0.05) based on molecular modules of the PD-I, PD-M, and PD-R, respectively.

Particularly, drug candidates predicted by targeting PD-R-specific gene module fell into fourteen pharmacological categories (according to Anatomical Therapeutic Chemical [ATC] code), including agents for the nervous and cardiovascular systems, immunomodulating agents, etc. The top-ranked drug candidates for PD-R were highlighted in Fig. 5a. Of note, our predictions included three US Food and Drug Administration (FDA)-approved drugs for PD treatment, including biperiden, amantadine, and levodopa. This demonstrated that our method is capable of predicting effective medication for PD. In addition, our data suggested some repurposable drug candidates. For instance, ambroxol^47^, an FDA-approved drug for respiratory disorders was predicted to be potentially useful for PD-R. Ambroxol has shown promise as a potential disease-modifying treatment for PD in a phase II, single-center, double-blind, randomized placebo-controlled trial^48^. Guanabenz, another drug prioritized for PD-R, is traditionally used for hypertension treatment. Guanabenz has been found to reduce 6-hydroxydopamine (6-OHDA)-induced cell death in ventral midbrain dopaminergic neurons in culture, and in dopaminergic neurons in the substantia nigra of mice.^49^

**Figure 5.**
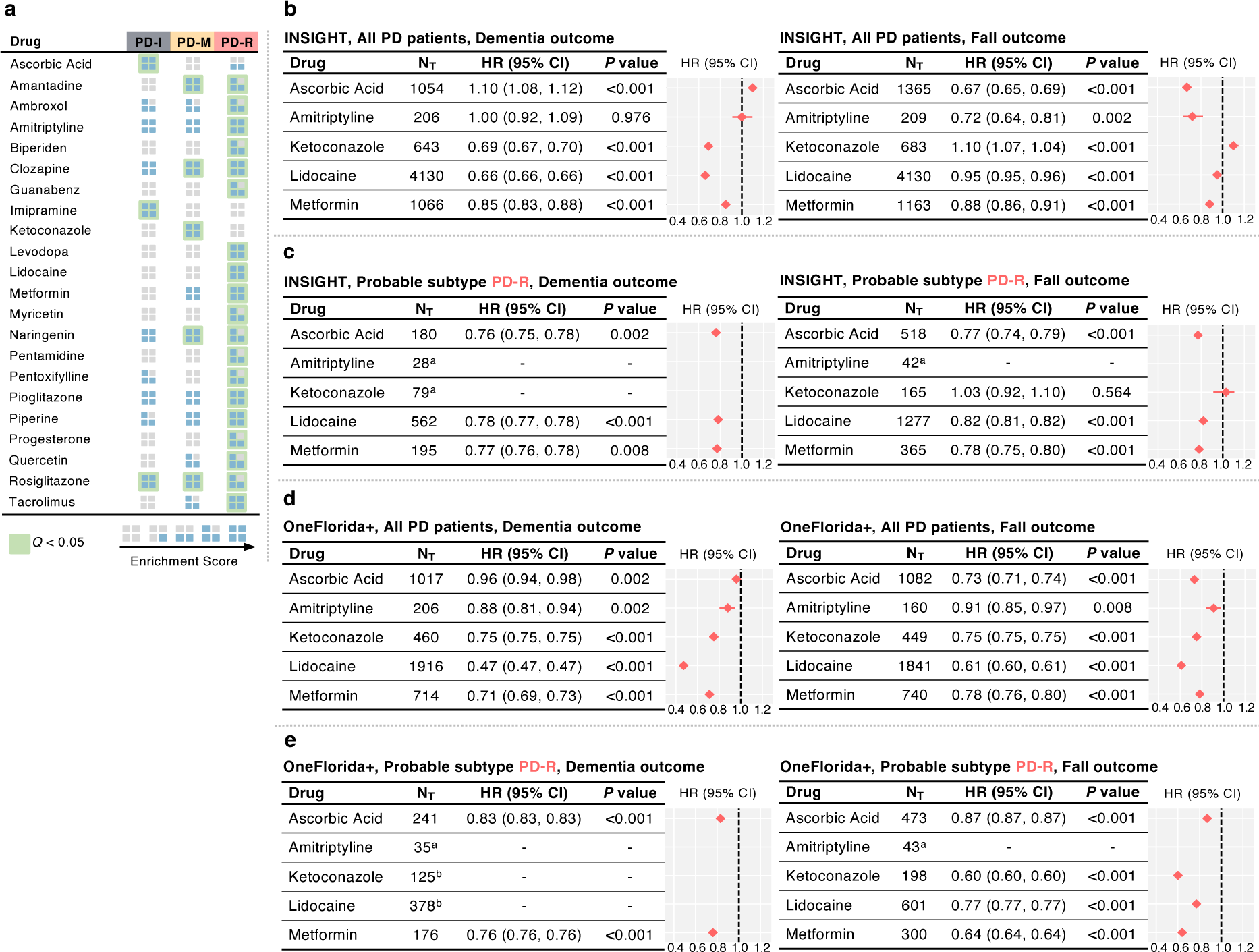
Identified repurposable drug candidates for preventing PD progression by targeting subtype-specific molecular changes. **a.** Gene set enrichment analysis (GSEA) informed by subtype gene modules with bulk RNA-seq data of individuals and transcriptomics-based drug-gene signature data in human cell lines identified repurposable drug candidates for different PD pace subtypes. **b-c.** Treatment effect estimation using the INSIGHT data within the broad PD population (**b**) and probable PD-R population (**c**). **d-e.** Treatment effect estimation using the OneFlorida+ data within the broad PD population (**d**) and probable PD-R population (**e**). ^a^The drug doesn’t have sufficient patient data (<100) for analysis. ^b^The drug doesn’t have sufficient balanced emulated trials (<10). N_T_ indicates the number of eligible PD patients who received the tested drug after PD initiation.

### Real-world evidence highlighted metformin as a promising candidate to mitigate PD progression

To gain RWE for validating treatment effects of the identified drug candidates, we used two large-scale, de-identified, patient-level RWD databases: the INSIGHT^50^, containing clinical information of >15 million patients in the New York City metropolitan area and the OneFlorida+^51^, covering ∼17 million patients in Florida, ∼2 million in Georgia, and ∼1 million in Alabama. We focused on five drugs based on subject matter expertise and a combination of multiple factors including: 1) the strength of network-based prediction (ES score > 3 and *Q* value < 0.05); 2) sufficient data for the target of interests; 3) pharmacokinetics profiles; and 4) knowledge functional data from the literature.

To evaluate treatment effects of the tested drugs, we conducted trial emulation based on our computational trial emulation framework^52^ (see Methods). Specifically, for each emulated trial, we built its treated group as the set of eligible patients who received a specific tested drug after PD initiation. Subsequently, we can formulate a control group as the set eligible patients who received alternative medications (i.e., those falling under the same ATC level 3 class as the tested drug, excluding the tested drug itself), through 1:1 nearest-neighbor matching. A propensity score method was used to learn empirical treatment assignment given the baseline covariates and an inverse probability of treatment weighting was used to balance the treated and control groups.^53^ We created 100 emulated trials for each tested drug and excluded drugs which had less than 10 balanced trials. We finally estimated drug treatment effects for the balanced trials using the Cox proportional hazard models.^54^ Following a previous study^55^, treatment efficiency was measured as the reduced risk to develop PD-related outcomes, including falls (a proxy of advanced motor impairment and dyskinesia relevant to PD progression) and dementia. More details of this analysis can be found in the Methods section and Supplementary Fig.3.

Using the eligible PD patient population within both INSIGHT and OneFlorida+ data, we were able to create sufficient (≥ 10) successfully balanced emulated trials for the five tested drugs. Metformin, an FDA-approved anti-diabetic medication, emerged as the most promising candidate for potentially preventing PD progression (see Fig. 5). Specifically, in the INSIGHT data, the usage of metformin was associated with 15% reduced likelihood of developing dementia (HR = 0.85, 95% CI [0.83, 0.88], *P* value < 0.001) as well as 12% reduced likelihood of onset of falls (HR = 0.88, 95% CI [0.86, 0.91], *P* value < 0.001), compared to the usage of alternative drugs (see Fig. 5b). Similarly, in the OneFlorida+ data, metformin usage was associated with 29% reduced likelihood of developing dementia (hazard ratio [HR] = 0.71, 95% CI [0.69, 0.73], *P* value < 0.001) as well as 22% reduced likelihood of onset of falls (HR = 0.78, 95% CI [0.76, 0.80], *P* value < 0.001) compared to the alternative medication usage (see Fig. 5d). Of note, we noticed a discrepancy in the therapeutic signals (i.e., HRs) of metformin between the INSIGHT and OneFlorida+ data. This could be attributed to variations in healthcare settings and population diversity within the two databases. However, what stands out more significantly is that our data, drawn from two independent real-world healthcare databases, consistently indicate that the use of metformin post-PD diagnosis may play a role in mitigating the progression of both motor and cognitive manifestations throughout the course of PD.

PD patients with early cognitive impairment are likely a subpopulation of PD-R (see Extended Data Fig. 1b). We further examined treatment efficiency of the drug candidates in this probable PD-R population. Specifically, we defined the probable PD-R as 1) patients diagnosed with PD, and 2) with any cognitive deficit diagnosis within the first year following PD initiation but prior to initiation of the tested drug. Under such settings, we repeated the trial emulation. We found that metformin could still be a good candidate for preventing PD progression within this population, and its treatment efficiency increased compared to that within the general PD population (see Fig. 5c). In the INSIGHT data, the usage of metformin was observed to be associated with 23% reduced likelihood of dementia (HR = 0.77, 95% CI [0.76, 0.78], *P* value <0.001) as well as associated with 22% reduced likelihood of fall events (HR = 0.78, 95% CI [0.75, 0.80], *P* value <0.001) compared to the non-metformin alternative drug usage (see Fig. 5c). Similarly, in the OneFlorida+ data, the usage of metformin was associated with 23% reduced likelihood of dementia (HR = 0.76, 95% CI [0.76, 0.76], *P* value <0.001) and with 36% reduced likelihood of fall events (HR = 0.64, 95% CI [0. 64, 0. 64], *P* value <0.001) compared to the alternative medication usage (see Fig. 5e)

## Discussion

The heterogeneous nature of PD progression has been widely acknowledged, yet it remains incompletely deciphered.^6–8^ In this investigation, we conducted comprehensive analyses of various types of data to study PD, including longitudinal clinical data, biospecimen, neuroimaging, genetic, and transcriptomic information of patients, publicly available PPI data and transcriptomics-based drug-gene signature data in cell lines (CMap), and two large-scale patient-level RWD databases (INSIGHT and OneFlorida+). We utilized cutting-edge data science approaches such as machine learning, deep learning, and network medicine techniques to unravel the progression heterogeneity of PD to identify PD subtypes; pinpoint subtype-specific biomarkers, genetic contributors, and molecular networks; and select potentially repurposable medications to ameliorate disease progression by targeting the subtypes.

The identification of clinical subtypes has garnered significant attention from researchers for better understanding PD heterogeneity.^8–11^ Conventional methods, which relied on a priori hypotheses focusing on single-domain information (e.g., tremor), have resulted in many clinical subtypes of PD. Notable examples included the motor subtypes such as tremor dominant (TD), akinetic-rigid (AR), and postural instability and gait disorder (PIGD). Moreover, recent data-driven methods have become increasingly popular in subtyping diseases such as PD, due to their hypothesis-free nature that allows unbiased investigation of vast and complex clinical data to identify subtypes. Compared to the previous PD subtyping systems, advantages of our PD pace subtypes are as follow (see Fig. 2b). First, our subtyping approach was completely data-driven and hypothesis-free. Second, recent cohort studies have demonstrated that the conventional motor subtypes were unstable over time.^12,13^ Such instability indicates that, these subtypes may be representative of various PD progression stages rather than distinct biological entities.^56^ Additionally, prior data-driven methods were mainly based on individual’s data collected at a specific time point (e.g., the baseline visit). This limited their capacities to accurately capture the evolving patterns of PD progression to address subtype infidelity over time. Our approach differs by leveraging deep learning to model 5-year high-dimensional, longitudinal clinical progression data of individuals. In this way, our approach captures the intrinsic phenotypic progression profiles of individuals to detect subtypes. Therefore, individuals’ subtype memberships were inherently stable over time, with each subtype reflecting a distinct PD progression pattern (in Fig. 2 and Table. 2).

A remarkable finding of us is the PD-R subtype, which had worse symptom severities at baseline and, more importantly, experienced the most rapid motor and non-motor symptom progression paces throughout the PD course. A recent data-driven study^57^ has reported a “diffuse malignant” clinical subtype of PD, which also exhibited poor baseline clinical manifestations and faster progression during follow-up. However, the progression speed of this “diffuse malignant” subtype was only marginally faster than the others and was noted in only a few clinical assessments, such as the Schwab and England score, MoCA, and global composite outcome. In contrast, PD-R demonstrated the greatest progression rates in a diverse spectrum of motor and non-motor manifestations among the PD population, estimated by the linear mixed effects models that considered multiple covariates (age, sex, levodopa equivalent dose [LED] usage at visits) (see Table 2 and Supplementary Table 8). Additionally, when compared to other subtypes, PD-R showed a more consistent trend to shift from TD toward PIGD (see Extended Data Fig. 1a). This not only strengthened the viewpoint that the TD and PIGD motor phenotypes may represent different stages of PD, but also verified that PD-R progressed faster than the others. Similarly, PD-R was more likely to develop cognitive and behavioral abnormalities, including MCI and/or dementia as well as depression (see Extended Data Fig. 1b-c).

Another two subtypes, i.e., PD-I and PD-M, exhibited comparable symptom severities at baseline (see Table 1). Nevertheless, importantly, substantial differences in clinical manifestations, both motor and non-motor, became apparent after two years from baseline (see Fig. 2a, and Supplementary Tables 3 and 4). PD-M manifested a much faster annual progression in most clinical manifestations compared to PD-I (see Table 2). The findings suggested that even when individuals with PD initially presented with similar symptom severities at baseline, their progression can vary in pace, leading to distinct trajectories and prognoses. This also supported the hypothesis that subtypes defined relying on baseline data can display fluctuations in symptom severity and/or outcomes during longitudinal follow-up.^12–14^ It underscores the importance of thoroughly modeling individual’s symptom progression trajectories when attempting to identify PD subtypes.

Given the distinct long-term (over 5 years) progression patterns of the PD subtypes, we were prompted to explore whether we could predict an individual’s subtype membership earlier. We identified a collection of CSF biomarkers, brain imaging markers, and genetic factors of the subtypes (see Fig. 3 and Extended Data Fig. 3), and we were able to construct a classification model of the subtypes. This model was based solely on a combination of demographic information, genetic factors, and clinical information collected within the first year after baseline including clinical assessments, CSF biomarkers, and brain imaging. The substantial performance (see Extended Data Fig. 10) demonstrated two key aspects of our model: 1) it amplified the prognostic value by enabling the identification of subtypes with distinct long-term progression patterns at an earlier stage (first year after baseline); 2) it facilitated the prediction of new patients’ subtype memberships, which, in turn, can accelerate precise patient management.

Cognitive impairment-related pathology may play a role in driving rapid PD progression, i.e., PD-R. Firstly, compared to the other subtypes, PD-R had worse cognitive performances in numerous clinical assessments and demonstrated an 8 to 10-fold higher prevalence of cognitive impairment at baseline (see Table 1 and Extended Data Fig. 1b). Secondly, the studied PD participants had no AD diagnosis at baseline and follow-up visits, yet our analysis of baseline CSF biomarkers suggested that several AD-type biomarkers could serve as potential early indicators for the distinct PD pace subtypes we identified (see Fig. 3a). More specifically, the CSF P-tau/*α*-synuclein ratio appeared to be a promising biomarker for differentiating the three PD subtypes from one another at baseline, although it had limited capacity to distinguish these subtypes against HC participants. Aside from the P-tau/*α*-synuclein ratio, we didn’t identify any additional CSF biomarkers that could effectively separate the PD-I and PD-M at baseline, which was consistent with the mixed symptom severities of the two subtypes at baseline. Other AD-related biomarkers including CSF A*β*-42, A*β*-42/P-tau ratio, and A*β*-42/T-tau ratio distinguished the PD-R from the PD-I and PD-M subtypes. Our findings are supported by the prior study^58^, which suggested that, although the AD biomarkers alone are not helpful in diagnosing PD, they could improve prognostic evaluation. This emphasizes the critical role that the primary hallmarks of AD, the amyloid and tau pathologies, may have in the progression of PD. Last, we also observed that early (within the first year after baseline) structural atrophy in certain brain regions responsible for cognitive functions may be potential markers of the PD pace subtypes (see Fig. 3b). Some notable markers we found included frontal lobe regions including rostral middle frontal, pars triangularis, caudal middle frontal, superior frontal gyri, as well as fusiform and parahippocampal gyri.

Analyses of individuals’ genetic and transcriptomic profiles armed with network medicine approaches led to the identification of subtype-specific molecular modules and biological pathways. Despite the commonalities in certain pathways across subtypes, each subtype was associated with unique pathways. This suggested that there might be unique pathophysiological underpinnings that drive the distinct progression patterns (i.e., the pace subtypes) observed in PD (see Extended Data Figs. 4e and 9c). In particular, the molecular modules specific to PD-R subtype suggested the potential roles of neuroinflammation, oxidative stress, metabolism, and AD pathways, along with PI3K/AKT and angiogenesis pathways in rapid PD progression (see Fig. 4). Neuroinflammation, a crucial factor in PD pathogenesis, involves the chronic inflammatory response in the brain, particularly the activation of microglia.^59^ Upon activation, microglia release pro-inflammatory cytokines and reactive oxygen species (ROS). Neuroinflammation-related ROS production subsequently leads to oxidative stress and neuronal damage, thereby playing a significant role in PD’s pathogenesis and progression.^59,60^ The PI3K/AKT pathway is fundamental to various physiological processes, such as cell survival, growth, proliferation, and metabolism. It also plays a crucial role in modulating the mammalian target of rapamycin (mTOR), a pivotal regulator of autophagy—an essential cellular process involved in degrading and recycling damaged or unnecessary cell components.^61,62^ Previous evidence has indicated that disruptions in the PI3K/AKT pathway impairs autophagy, leading to ineffective elimination of abnormal and toxic protein aggregates, contributing to neuronal death. ^57,58^ In addition, angiogenesis, the process of forming new blood vessels from existing ones, has been implicated in PD pathologies.^63^ Aberrant angiogenesis has been associated with blood-brain barrier (BBB) dysfunction. This dysfunction may lead to changes in BBB permeability, potentially permitting neurotoxic substances to enter the brain.^64^ It could also trigger neuroinflammation and abnormal immune responses, both contributing to neurodegeneration.^64^

Using in-silico drug repurposing approaches, we prioritized drug candidates that could target specific PD subtypes (see Fig. 5a). Metformin, a traditional type 2 diabetes (T2D) medication, stood out as a potentially promising candidate for repurposing. Our data demonstrated the potential effect of metformin in counteracting molecular changes triggered in PD-R (ES = 4.44, *P* value < 0.001). Furthermore, real-world evidence gained from large-scale individual-level RWD substantiated our findings, showing that metformin usage was associated with a decreased risk of advanced PD outcomes including dementia (cognitive impairment) and falls (advanced motor dysfunction). Notably, a stronger treatment effect was observed within the surrogate PD-R subtype population (INSIGHT data: dementia, HR = 0.77, 95% CI [0.76, 0.78], *P* = 0.008; falls, HR = 0.78, 95% CI [0.75, 0.80], *P* value < 0.001; OneFlorida+: dementia, HR = 0.71, 95% CI [0.69, 0.73], *P* value < 0.001; falls, HR = 0.78, 95% CI [0.76, 0.80], *P* value < 0.001) compared to that within the broader PD population (INSIGHT data: dementia, HR = 0.85, 95% CI [0.83, 0.88], *P* value < 0.001; falls, HR = 0.88, 95% CI [0.86, 0.91], *P* value < 0.001; OneFlorida+: dementia, HR = 0.76, 95% CI [0.76, 0.76], *P* value <0.001; falls, HR = 0.64, 95% CI [0. 64, 0. 64], *P* value <0.001). Metformin, known for its efficacy in reducing hepatic glucose release and augmenting the body’s insulin sensitivity, has recently also been highlighted for its potential neuroprotective role in neurodegenerative disorders including PD. This is supported by a growing body of *in vitro* and *in vivo* studies of PD.^65–67^ First, known as a strong AMPK (5’-adenosine mono-phosphate-activated protein kinase) activator, metformin’s neuroprotective effects may be primarily attributable to its role in activating the AMPK signaling pathway.^68,69^ Past research found that AMPK can suppress microglial activation, thus potentially mitigating neuroinflammation.^68^ Recent investigations employing animal models revealed metformin’s ability to reduce microglial cell numbers, modify microglial activation pathways, and attenuate both inflammasome activation and the accumulation of ROS in microglia.^70,71^ In addition, metformin has been found to modulate PI3K/AKT activity.^72,73^ Enhanced activations of AMPK and PI3K/AKT can amplify autophagy, which aids in the removal of toxic proteins like *α* -synuclein, thereby preventing dopaminergic neuronal death.^72,73^ Finally, animal model studies have found that metformin may induce both angiogenesis and neurogenesis in the brain under different experimental conditions.^74–76^ In conclusion, the neuroprotective effect of metformin against the PD-R subtype (rapid PD progression) could be attributed to its ability to inhibit neuroinflammation, enhance PI3K/AKT activity for efficient *α* - synuclein clearance, and stimulate angiogenesis. Our data further supported the potential of metformin in treating PD, especially the rapid pace PD subtype patients, but also indicated the potential of our method in accelerating precision medicine approaches towards identifying therapeutics that reduce PD progression.

Limitations of this work should also be acknowledged. First, the subtypes were detected and validated based on the PPMI and PDBP cohorts. Though the two cohorts provided excellent phenotypic, molecular, and imaging characterization, they included highly selected (biased) samples (high education level and more white people). This could limit the generalizability of our findings and further validation in cohorts representing more diverse population is needed. Second, data incompleteness could impact the subtype findings. To address this issue, we excluded individuals who had insufficient longitudinal information and used a combination of the last observation carried forward (LOCF) and next observation carried backward (NOCB) methods^77^ to impute missing values while retaining longitudinal consistency. We also imputed the variables missing all values along the timeline of an individual based on median values of the population. Even so, we may have failed to reflect the real values and thus introduced noise and bias in analysis. Third, we investigated transcriptomic profiles from whole blood samples. Given the distinct roles different tissues play in PD, comprehensive investigations integrating other tissue data and additional omics data – such as proteomics and single-cell RNA-seq – are needed to uncover more accurate biological underpinnings of the identified subtypes. Last, translating subtype findings from research cohorts (e.g., PPMI and PDBP) to RWD to gather real-world evidence necessarily posed significant challenges for application to the general population due to selection bias in research cohorts. We defined surrogate PD-R in RWD based on its unique clinical feature of early cognitive impairment, which may be not accurate. Future research employing more sophisticated techniques like transfer learning, which facilitates the transfer of knowledge across cohorts with partially overlapping features is needed. In addition, RWD analysis may not be suitable for testing drugs that have too few patient data. We were able to perform population-based validation for metformin, as it had a large amount of patient data available. For drugs with fewer patient data, replication of the associations using multiple large population-based cohorts is suggested. Last, our RWD analysis cannot build causal relationships between metformin use and beneficial clinical response of PD or PD-R. Causal methods, such as Mendelian randomization studies^78,79^, should be utilized in the future.

## Summary

Through the integrative analyses of multi-modal clinical progression data, CSF biomarkers, brain imaging, and multi-omics data, we identified PD subtypes with a focus on clarifying clinical progression heterogeneity of PD, as well as pinpointing subtype-specific biomarkers, genetic contributors, and molecular networks. This established the foundation for a deeper understanding of biological mechanisms that drive the heterogeneity in PD progression. By integrating in-silico drug repurposing approaches with large-scale patient RWD, we identified metformin as a potential treatment for slowing PD progression. The findings deepen our understanding of the pathobiology of this complex disease and move the field toward precision medicine.

## Methods

### Study cohorts for PD subtyping

The present study included two longitudinal PD cohorts for identifying PD subtypes: the Parkinson’s Progression Markers Initiative (PPMI, http://www.ppmi-info.org)^25^ and the Parkinson Disease Biomarkers Program (PDBP, https://pdbp.ninds.nih.gov)^26^.

PPMI, launched in 2010 and sponsored by the Michael J. Fox Foundation, is an international and multi-center observational study dedicated to identifying biomarkers indicative of PD progression^25^. The present study included the following participants in the PPMI cohorts: *de novo* PD participants (diagnosed with PD within the last 2 years and untreated at enrollment), HCs, and individuals with SWEDD (scans without evidence of dopaminergic deficit). More information about PPMI participants has been described elsewhere^25^. The PPMI study protocol was approved by the institutional review board of the University of Rochester (NY, USA), as well as from each PPMI participating site. Data were obtained from PPMI website (https://www.ppmi-info.org) under PPMI Data Use Agreement.

PDBP, established in 2012 and funded by the National Institute of Neurological Disorders and Stroke (NINDS), is an observational study for advancing comprehensive PD biomarker research^26^. The present study included the participants with PD and HCs in the PDBP cohort. More information about the PDBP participants has been described elsewhere^26^. The study protocol for each PDBP site was approved by institutional review board of each participating site. Data were obtained via the Accelerating Medicines Partnership Parkinson’s disease (AMP-PD) platform (http://amp-pd.org) under AMP-PD Data Use Agreement.

We used the PPMI cohort as the development cohort. Participants who had less than 1-year historical records were excluded as lack of longitudinal information. Participants’ longitudinal clinical data were collected for modeling PD symptom progression trajectories to produce a progression embedding vector for each participant, using deep learning. Subtypes were identified using the learned progression embedding vectors of participants with PD. We used the PDBP cohort as the validation cohort. Participants who had less than 1-year historical records were excluded. In the PDBP, only the early PDs (symptom duration < 3 years at enrollment) were used to re-identify the PD subtypes. More details of the studied cohorts and data utilization were illustrated in the Supplementary Table 1. All study participants provided written informed consent for their participation in both studies.

### PD progression modeling for subtype identification

#### Clinical variables

We used participant’s longitudinal data in diverse clinical assessments. Specifically, we used motor manifestation data including Movement Disorders Society–revised Unified Parkinson’s Disease Rating Scale (MDS-UPDRS) Parts II and III^80^ and Schwab-England activities of daily living score, as well as non-motor manifestation data including MDS-UPDRS Part I^80^, Scales for Outcomes in Parkinson’s disease-Autonomic (SCOPA-AUT)^81^, Geriatric depression scale (GDS)^82^, Questionnaire for Impulsive-Compulsive Disorders in Parkinson’s disease (QUIP)^83^, State-Trait Anxiety Inventory (STAI)^84^, Benton Judgment of Line Orientation (JOLO)^85^, Hopkins Verbal Learning Test (HVLT)^86^, Letter-number sequencing (LNS), Montreal Cognitive Assessment (MoCA)^87^, Semantic verbal-language fluency test^88^, Symbol–Digit Matching (SDM)^89^, Epworth Sleepiness Score (ESS)^90^, REM sleep behaviour disorder (RBD)^91^, and Cranial Nerve Examination. Usage of dopaminergic medication was transformed into levodopa equivalent daily dose usage.^92^

For the PDBP cohort, we used the clinical variables shared with the PPMI cohort. More details of the clinical variables used for PD subtyping can be found in the Supplementary Table 2.

#### Data preparation

In both the PPMI and PDBP cohorts, we extracted clinical data at baseline and follow-up visits for each participant. In this way, each participant was associated with a multivariate clinical time sequence. Missing values were imputed using the combination of last observation carried forward (LOCF) and next observation carried backward (NOCB) strategies which has demonstrated effectiveness in our previous work^77^. The variables missing all values along timeline of a participant were imputed by median values of the population. To eliminate the effects of value magnitude, all variables were scaled based on z-score.

#### Learning individuals’ phenotypic progression embedding vectors using deep learning

Our goal was to identify PD subtypes, each of which can reflect a unique PD symptom progression pattern within the course of PD. To this end, there is the need of fully considering longitudinal data of individuals to derive PD subtypes. Here, we developed a deep learning model, termed deep phenotypic progression embedding (DPPE), which took the multivariate clinical time sequence data of each participant as input to learn a machine-readable vector representation, encoding his/her PD phenotypic progression trajectory over time (see Fig. 1g). Specifically, the DPPE model was based on the Long-Short Term Memory (LSTM) units^28,29^, a deep learning model designed for time sequence data modeling. The LSTM unit has a “memory cell” that stores historical information for extended periods, making it an excellent choice for modeling disease progression using longitudinal clinical data.^93^ The DPPE engaged an autoencoder architecture that consisted of two components, an encoder and a decoder, each of which is a LSTM model: 1) the encoder took the longitudinal clinical data of each individual (i.e., multivariate time sequence) as input and learned a progression embedding vector that encoded his/her PD symptom progression trajectory; and 2) the decoder, with reversed architecture of the encoder, tried to reconstruct the input time sequence of each individual based on the embedding vector. The DPPE model was trained by minimizing difference between the input multivariate clinical time sequences and the reconstructed ones. To enhance model training, we used data of PDs, HCs, and SWEDDs in the PPMI cohorts. After training of the DPPE model, we obtained a learned progression embedding vector for each participant, encoding his/her PD progression profile.

#### Cluster analysis for subtype identification in the PPMI cohort

The agglomerative hierarchical clustering (AHC) with Euclidean distance calculation and Ward linkage criterion^30^ was applied to the individuals’ embedding vectors learned by the DPPE model. We used AHC because that, 1) unlike other clustering methods like k-means clustering (which requires a sphere-like distribution of the data), AHC is usually robust as it’s not sensitive to the distribution of the data and doesn’t require an initialization procedure (e.g., k-means) that may incorporate uncertainty; 2) AHC can produce a tree diagram known as a dendrogram, visually interpreting how the data points are agglomerated together in a hierarchical manner and also illustrating the distances between the clusters at different layers in the hierarchy, providing visible guidance in determining the optimal cluster number. AHC has previously shown promise in identifying underlying patterns from clinical profiles for disease subtyping.^94–96^

A crucial issue of cluster analysis is the determination of cluster number in data. To address this issue, we considered multiple criteria to determine the optimal cluster (i.e., subtype) number in cluster analysis based on AHC: 1) Clusters should be clearly separated in the dendrogram produced by the AHC. 2) We selected optimal cluster numbers suggested based on clustering measurements calculated by the ‘NbClust’ software^31^, an R package for assisting a clustering method to determine the optimal cluster number of the data. Here, we used 18 indices provided by ‘NbClust’ to evaluate cluster structure of the agglomerative hierarchical clustering model with Ward criterion. The optimal cluster number was determined by the optimal value of each index. The used indices included: Scott index, Marriot index, TrCovW index, TraceW index, Friedman index, Rubin index, DB index, Silhouette index, Duda index, Pseudot2 index, Beale index, Ratkowsky index, Ball index, Ptbiserial index, Frey index, McClain index, Dunn index, SDindex. More details of these indices were introduced elsewhere^31^. 3) We calculated the 2-dimensional (2D) representation for each individual based on his/her progression embedding vector using the t-distributed stochastic neighbor embedding (t-SNE) algorithm^32^. We then visualized individuals’ subtype memberships in the 2D t-SNE space. We expected that the clusters, i.e., PD subtypes, could be clearly separated in the 2D t-SNE space. 4) We also considered clinical interpretations of the subtype results to determine the optimal cluster number.

#### Subtype validation in the PDBP cohort

To enhance reproducibility of the identified subtypes, we validated them using the PDBP cohort. Specifically, we repeated the whole procedure above in the PPMI cohort to re-identify the subtypes in the PDBP cohort. We re-trained the DPPE model to calculate individuals’ progression embedding vectors. We used data of HCs and all participants with PD in the PDBP cohort for training the DPPE model. PD participants in the PDBP cohort had a broad distribution of PD duration^26^. To align with our primary analysis in the PPMI cohort, we used the individuals with early PD (whose PD duration < 3 years) and performed AHC based on their learned embedding vectors to re-derive subtypes.

### Exploring clinical characteristics of the identified subtypes

We characterized the identified subtypes in two ways. First, we characterized the subtypes by evaluating their differences in demographics and clinical assessments at baseline as well as 2- and 5-year follow-up. For group comparisons, we performed analysis of variance (ANOVA) for continuous data and *χ*^2^ test for categorical data. Analysis of covariance (ANCOVA) was also applied, adjusting for age, sex, and levodopa equivalent dose (LED) usage. A two-tailed *P* value < 0.05 were considered as the threshold for statistical significance.

Second, we estimated annual progression rates in terms of each clinical assessment for each subtype. To accomplish this, for each variable, we fitted a linear mixed effect model for each PD subtype, specifying time (year) as the explanatory variable of interest. For all models, individual variation was included as a random effect, and age, sex, and LED at visits were included as the covariates. For each model, we reported the coefficient *β* (95% CI) of time as annual progression rate of the specific clinical assessment, along with the corresponding *P* value. A *P* value <0.05 was considered for statistical significance. We further utilized Sankey diagrams to visualize the transition trends of motor phenotypes (tremor dominant and postural instability and gait disorder [PIGD]) as well as non-motor phenotypes including cognition (normal cognition, mild cognition impairment [MCI], and dementia), REM sleep behavior disorder (RBD negative or positive), and depression (normal, as well as mild, moderate, and severe depression).

### CSF biomarker analysis

CSF biospecimen data were obtained from the PPMI cohort. We used baseline *α*-synuclein measured by enzyme-linked immunosorbent assay^97^, amyloid-beta1–42 (*Aβ*-42), phosphorylated Tau protein at threonine 181 (P-tau), and total tau protein (T-tau) measured by INNO-BIA AlzBio3 immunoassay. Following the previous studies^98,99^, we also evaluated *Aβ*-42/P-tau, *Aβ*-42/T-tau, *Aβ*-42/*α*-synuclein, P-tau/*α*-synuclein, T-tau/*α*-synuclein, and P-tau/T-tau levels. We conducted two-group comparisons (subtype vs. subtype and subtype vs. HCs) for each biomarker using linear mixed effect models, specifying individuals’ HC or PD subtype membership as the explanatory variable of interest, adjusting for baseline age and sex as covariates. A *P* value < 0.05 was considered for statistical significance. Additionally, boxplots were engaged to visualize data distributions.

### Neuroimaging biomarker analysis

High resolution T1-weighted 3 T Magnetic Resonance Imaging (MRI) data of participants were available at baseline and follow-up in the PPMI cohort. For each individual, we calculated 1-year brain atrophy measured by cortical thickness and white matter volume in 34 brain regions of interests (ROIs), defined by the Desikan-Killiany atlas (averaged over the left and right hemispheres). We evaluated those measures in separating each pair of PD subtypes. The student’s t-tests were used for two-group comparisons. The ‘ggseg’^100^ in R was used to visualize the neuroimaging biomarkers under the Desikan-Killiany atlas.

### Construction of human protein-protein interactome network

We assembled commonly used human protein-protein interactome (PPI) databases with experimental evidence and in-house systematic human PPIs to build a comprehensive human PPI network. PPI databases we used included: (i) kinase-substrate interactions via literature-derived low-throughput and high-throughput experiments from Human Protein Resource Database (HPRD)^101^, Phospho.ELM^102^, KinomeNetworkX^103^, PhosphoNetworks^104^, PhosphositePlus^105^, and DbPTM 3.0^106^; (ii) binary PPIs from 3D protein structures from Instruct^107^; (iii) binary PPIs assessed by high-throughput yeast-two-hybrid (Y2H) experiments^108^; (iv) protein complex data (∼56,000 candidate interactions) identified by a robust affinity purification-mass spectrometry collected from BioPlex V2.0^109^; (v) signaling networks by literature-derived low-throughput experiments from the SignaLink2.0^110^; and (vi) literature-curated PPIs identified by affinity purification followed by mass spectrometry from BioGRID^111^, HPRD^112^, InnateDB^112^, IntAct^113^, MINT^114^, and PINA^115^. In total, 351,444 PPIs connecting 17,706 unique proteins are now freely available at https://alzgps.lerner.ccf.org.^116^ In this study, we utilized the largest connected component of this dataset, including 17,456 proteins and 336,549 PPIs. The PPI network was used for network analyses of genetic and transcriptomic data for subtype-specific molecular module identification, which were detailed below.

### Genetic data analysis

To explore genetic components of the PD subtypes, we analyzed genetic data in the PPMI cohort. APOE *ε* 2 and *ε* 4 genotypes and genotypes of 90 PD-related risk loci^33^ were collected for analysis. The hypergeometric tests were used to identify SNPs that were enriched in each subtype within the PD population. *P* value from the hypergeometric analysis indicates association of a SNP to the specific subtype^34^. A *P* value<0.05 was considered for statistical significance.

### Construction of genetic molecular modules of the identified subtypes

Network analysis was conducted based on the PPI network we built to expand the genetic signals to identify subtype-specific molecular module. For each subtype, we first selected the SNPs that were associated with the subtype (*P* value < 0.05) and obtained their nearest genes and/or possible causal genes identified through the PD GWAS Locus Browser^117^. The PD GWAS Locus Browser identified causal genes for each PD risk loci, as protein coding genes within 1 Mb up and downstream of it, via integration of diverse data resources, such as gene expression and expression quantitative trait locus (eQTL) data from different tissues as well as literature. This resulted in a list of genetic associated genes for each subtype. On the other hand, we conducted an analysis of single-nucleus RNA (snRNA) sequencing data from brain tissues to identify a set of PD contextual genes (see Supplementary Methods). Then, for each subtype, we linked its genetic associated genes with the PD contextual genes in the PPI network we built. Genetic associated genes that cannot link to any contextual genes were removed. In this way, these linked genetic associated genes and their linked contextual genes constructed the genetic molecular module specific to the subtype.

### Transcriptomic data analysis

We performed gene expression analysis using whole blood bulk RNA-seq data of participants in the PPMI cohort^118^. Genes were annotated using UniProt^119^, and only protein coding genes were included for analysis. Genes with low expression levels across all samples were excluded. Differential gene expression analyses were performed for each subtype compared to healthy controls using DEseq2^120^. Age, sex, and LED usage were included as covariates. An adjusted *P* value<0.05 was considered for statistical significance.

### Construction of transcriptomic molecular modules of the identified subtypes using GPSnet algorithm

Our GPSnet (Genome-wide Positioning Systems network)^38^ demanded two inputs: the node (gene) scores and a background PPI network we built above. GPSnet first set an initial gene score *z*(*i*) for each gene *i* in the PPI network: for differentially expressed genes (DEGs) with *Q* value ≤ 0.05, the gene scores were initialized as *z*(*i*) = |*log*_2_FC|, where FC indicates fold change; for the remaining non-DEGs, *z*(*i*) = 0. After that, a random walk with restart process was applied to smoothen the scores for all genes in the PPI network. Next, GPSnet utilized the following procedures to build a gene module *M*: It first initialized module *M* only containing a randomly selected seed gene and then expanded the module by involving genes one by one, while 1) enhancing gene connectivity within the module and 2) improving module level gene score. Specifically, a gene *i* ∈ *Γ_M_* will be included into *M* (*Γ_M_* is a set of genes that interact with genes in module *M*), if 1) *P*(*i*) ≤ 0.01 (calculated with **Eq. (1)**, indicating genes within *M* densely connect to each other after including *i*) and 2) *S*(*M* ∪ {*i*}) > *S*(*M*) (calculated with **Eq. (2)**, indicating module-level risk score increased after involving gene *i*).

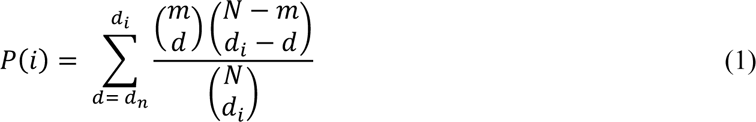

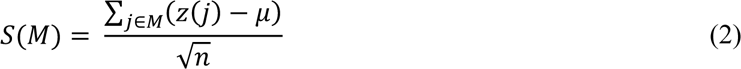

where, *m* denotes the number of genes in module *M*, *μ* denotes the average score of all genes in the PPI network, and *S*(*M*) denotes the module-level gene score of the module *M*.

We stopped module expansion when *S*(*M*) could or be increased or *P*(*i*) ≥ 0.01 by involving new genes. In this study, we repeated above procedures multiple (∼100,000) times and obtained a collection of raw modules. All raw modules were ranked in a descending order based on module score. We generated the final gene modules by assembling the top ranked modules. For each subtype, we ran GPSnet based on the DEGs of a specific subtype to gain the transcriptomic molecular module of the subtype.

### Pathway enrichment analysis

Pathway enrichment analyses were conducted based on using BioPlanet^121^ from Enrichr^122^. For each subtype, we identified pathways according to the genetic and transcriptomic molecular modules of the subtype, respectively. The combined score defined in Enrichr^122^ equaled the product of log of *P* value from the Fisher’s test and z-score which characterized the departure from the expected rank.

### Building classification model of subtypes

To gain more prognostic insights of the subtypes, we built a prognostic model of subtypes using information collected within the first year after enrollment. Specifically, we used candidate predictors including demographics, 10 principal components derived from neuroimaging biomarkers (1-year brain atrophy measured by reduction of cortical thickness and white matter volume in 34 brain ROIs), 90 PD-related SNPs, and clinical variables at baseline and 1-year follow-up. To improve model practicability in separating multiple subtypes, we built the model using a cascade framework^44^, which was proposed to address multi-label classification tasks. Specifically, it contained a sequential of multiple basic classifiers, such that in each step, it predicted a subtype from the remaining subjects that will be sent to the successor basic classifier. We used random forest as the basic classifier and the model was trained using a 5-fold cross-validation strategy. We used the receiver operating characteristics curve (ROC) and area under ROC curve (AUC) to measure prediction performance of our model.

### Subtype-specific in-silico drug repurposing

#### Connectivity Map (CMap) database

We downloaded transcriptomics-based drug-gene signature data in human cell lines from the Connectivity Map (Cmap) database^24^. The Cmap data used in this study contained 6,100 expression profiles relating 1,309 compounds^24^. The CMap provided a measure of the extent of differential expression for a given probe set. The amplitude *α* was defined as **Eq. (3)** as follows:

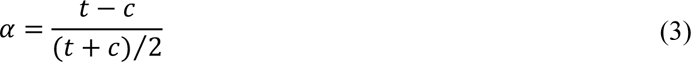

where *t* is the scaled and threshold average difference value for the drug treatment group and *c* is the threshold average difference value for the control group. Therefore, an *α*=0 indicates no expression change after drug treatment, while an *α* > 0 indicates elevated expression level after drug treatment and vice versa.

#### Gene set enrichment analysis (GSEA) for drug repurposing

We applied GSEA algorithm for predicting repurposable drug candidates for each PD subtype. The GSEA algorithm demanded two inputs: the CMap data and a list of module genes. For each subtype, we combined the genetic and transcriptomic molecular module to obtain a list of module genes for this subtype. Detailed descriptions of GSEA have been illustrated in elsewhere^38,46^. Then, for each drug in the CMap database, we calculated an enrichment score (ES) based on genes within each subtype-specific molecular module as **Eq. (4)**. ES represents drug potential capability to reverse the expression of the input molecular network:

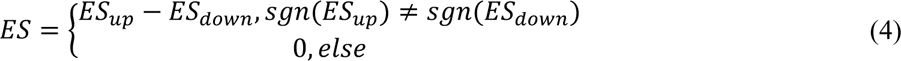

where *ES_up_* and *ES_down_* were calculated separately for up- and down-regulated genes from the subtype-specific gene module. We computed *a_up/down_* and *b_up/down_* as

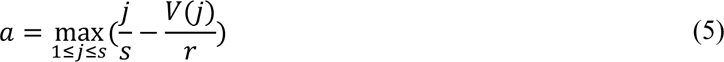

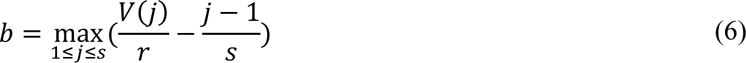

where *j* = 1, 2, 3, …, *s* are the genes within the subtype-specific module sorted in an ascending order by their rank in the gene expression profiles of the tested drug. The rank of gene *j* was defined as *V*(*j*), such that 0 ≤ *V*(*j*) ≤ *r* with *r* being the number of module genes in drug profile. Then *ES_up/down_* was set to *a_up/down_* if *a_up/down_* > *b_up/down_* and was set to −*b_up/down_* if *a_up/down_* < *b_up/down_* Permutation tests repeated 100 times using randomly selected gene lists consisting of the same numbers of up- and down-regulated genes as the input subtype-specific gene module were performed to calculate the significance of the computed ES value. Therefore, drugs with large positive ES values and *P* values ≤ 0.05 were selected.

### Pharmacoepidemiologic validation with large-scale real-world patient data

We estimated treatment effects of the identified repurposable drug candidates. Overall workflow of the real-world patient data analysis can be found in the Supplementary Fig. 3. The detailed procedures were introduced as below.

#### Real-world patient data

In this study, we used two independent large-scale real-world patient databases.

○ *<u>INSIGHT Clinical Research Network (CRN).</u>* The INSIGHT CRN^50^ was founded with and continues to be supported by Patient-Centered Outcomes Research Institute (PCORI). The INSIGHT CRN brings together top academic medical centers located in New York City (NYC), including Albert Einstein School of Medicine/Montefiore Medical Center, Columbia University and Weill Cornell Medicine/New York-Presbyterian Hospital, lcahn School of Medicine/Mount Sinai Health System, and New York University School of Medicine/Langone Medical Center. This study used de-identified patient real-world data (RWD) from the INSIGHT CRN, which contained longitudinal clinical data of over 15 million patients in the NYC metropolitan area. The use of the INSIGHT data was approved by the Institutional Review Board (IRB) of Weill Cornell Medicine under protocol 21-07023759.
○ *<u>OneFlorida+ Clinical Research Consortium.</u>* OneFlorida+ Clinical Research Consortium^51^ is another CRN supported by PCORI, which includes 12 healthcare organizations and contains longitudinal and linked patient-level data. This study used de-identified, robust linked patient-level RWD of 17 million patients in Florida, 2.1 million in Georgia (via Emory), and 1.1 million in Alabama (via UAB Medicine) since 2012 and covering a wide range of patient characteristics including demographics, diagnoses, medications, procedures, vital signs, and lab tests. The use of the OneFlorida+ approved by the IRB of University of Florida under protocol IRB202300639.

#### Eligibility criteria

Patient eligibility criteria for analysis included:

○ Patients should have at least one PD diagnosis according to International Classification of Diseases 9th and 10th revision (ICD-9/10) codes, including 332.0 (ICD-9) and G20 (ICD-10).
○ Patient’s age was >= 50 years old at the first PD diagnosis.
○ Patients who had neurodegenerative disease diagnoses before his/her first PD diagnosis was excluded.

#### PD outcomes

We considered PD related outcomes including dementia and falls (indicating advanced motor impairment and dyskinesia), which were defined by ICD-9/10 diagnosis codes. Drug treatment efficiency was defined by reducing the risk to develop the PD outcomes.

#### Follow-up

Each patient was followed from his/her baseline until the day of the first PD outcome event, or loss to follow-up (censoring), whichever happened first.

#### Trial Emulation

We first obtained DrugBank ID of each tested drug and translated it to RxNorm and NDC codes using the RxNav API (application programming interface). Drugs which were used by less than 100 patients were excluded for analysis. Following Ozery-Flato et al.^55^, we defined the PD initiation date of each patient as six months prior to his/her first recorded PD diagnosis event. This accounted for the likelihood that PD may be latently present before formal diagnosis. We defined the index date as the beginning time of treatment of a tested drug candidate or its alternative treatment. We also defined the baseline period as the time interval between the PD initiation date and the index date for each patient. We required that:

○ The index date was later than the PD initiation date.
○ Onset of PD outcomes were later than the index date.

Then, for each tested drug, we built an emulated trial using the following procedures:

1. We built its treated group as the eligible PD patients who took the tested drug after PD initiation;
2. We built a control group as:

- Patients who received alternative treatment of the tested drug, i.e., drugs from a same Anatomical Therapeutic Chemical level 3 [ATC-L3] classification of the tested drug, excluding the drug itself.
- To control confounding factors, we performed the propensity score matching as introduced below.

#### Propensity score matching (PSM)

We collected three types of covariates at the baseline period for each patient: 1) We included 64 comorbidities including comorbidities from Chronic Conditions Data Warehouse and other risk factors that were selected by experts.^123^ The comorbidities were defined by a set of ICD-9/10 codes. 2) We considered usage of 200 most prevalent prescribed drug ingredients as covariates in this analysis. These drugs were coded using RxNorm and grouped into major active ingredients using Unified Medical Language System. 3) We also included other covariates, including age, gender, race, and the time from the PD initiation date to the drug index date. In total, we included 267 covariates for analysis.

For each emulated trial, we used a propensity score framework to learn the empirical treatment assignment given the baseline covariates and used an inverse probability of treatment weighting to balance the treated and control groups.^53^ For each trial, a 1:1 nearest-neighbor matching was performed to build the matched control group.^53^ The covariate balance after propensity score matching was assessed using the absolute standardized mean difference (SMD).^124^ For each covariate, it was considered balanced if its SMD≤0.2, and the treated and control group were balanced if only no more than 2% covariates were not balanced.^125^ To enhance robustness of the analysis, we created 100 emulated trials for each tested drug. Tested drugs that had <10 successfully balanced trials were excluded for analysis.^53^

#### Treatment effect estimation

For each tested drug, we estimated drug treatment effect for each balanced trial by calculating the hazard ratio (HR) using a Cox proportional hazard model^54^, comparing the risk to develop a specific outcome between the treated and control groups. We reported the median HR with 95% confidence intervals (CI) obtained by bootstrapping.^126^ A hazard ratio < 1 indicated the tested drug can reduce risk to develop a specific outcome and a P value < 0.05 was considered as statistically significant.

## Data availability

Data from the PPMI can be download from the official web site (http://www.ppmi-info.org) via request. Data from the PDBP can be download from AMP-PD platform (http://amp-pd.org) via request. Information of the INSIGHT database is available at https://insightcrn.org. Request of INSIGHT data can be sent via: https://nyc-cdrn.atlassian.net/servicedesk/customer/portal/2/group/6/create/16. Information of the OneFlorida+ data is available at: https://onefloridaconsortium.org/. Request of OneFlorida+ data can be sent via: https://onefloridaconsortium.org/front-door/prep-to-research-data-query/.

## Code availability

Computer codes for this study are available at https://github.com/changsu10/Parkinson-Progression-Subtyping/tree/main.

## Author contributions

F.W. and C.S. conceived the study. F.W. secured funding and supervised the research. C.S. preprocessed clinical data and implemented subtyping model for primary analysis. Y.H. conducted subtype validation analysis. C.S. and Y.H. built subtype prediction model. C.S. and Y.H. conducted genetic data analysis. C.S., J.R.M., Z.B., and H.Z. conducted bulk transcriptomic analysis. X.S. advised genetic and transcriptomic analyses. Y.Z. advised imaging processing. C.S., and M.B. conducted imaging analysis. F.C. designed network medicine method and supervised network analysis and in-silico drug repurposing. J.-L.X. conducted network analysis and in-silico drug repurposing. J.B. supervised real-world data collection and analysis. Z.X. and J.X. conducted real-world data analysis. C.H., J.B.L., J.C., and M.C.C provided clinical expertise, reviewed the manuscript, and approved the final version. C.S. drafted manuscript. All authors critically revised the manuscript for important intellectual content. All authors provided comments and approved the paper.

## Acknowledgements

We thank the AMP-PD consortium for developing and providing access to the data resource. We thank the PPMI and PDBP cohorts for contributing their data to AMP-PD. This work was supported by following: National Institute on Aging RF1AG072449 and the Michael J. Fox Foundation to F.W.; National Institute on Aging R01AG080991 and R01AG076234 to F.W. and J.B.; National Institute on Aging R01AG076448 to F.W. and F.C.; National Institute on Aging U01AG073323, R21AG083003, R01AG066707, R01AG082118, RF1AG082211, RF1NS133812, 3R01AG066707-01S1, 3R01AG066707-02S1, and R56AG074001 and Alzheimer’s Association (ALZDISCOVERY-1051936) to F.C.; National Institute of General Medical Sciences P20GM109025, National Institute of Neurological Disorders and Stroke U01NS093334, National Institute on Aging R01AG053798, P30AG072959, R35AG71476, and AG083721-01, Alzheimer’s Disease Drug Discovery Foundation (ADDF), Ted and Maria Quirk Endowment, and Joy Chambers-Grundy Endowment to J.C..

## Extended data figures

**Extended Data Fig. 1.**
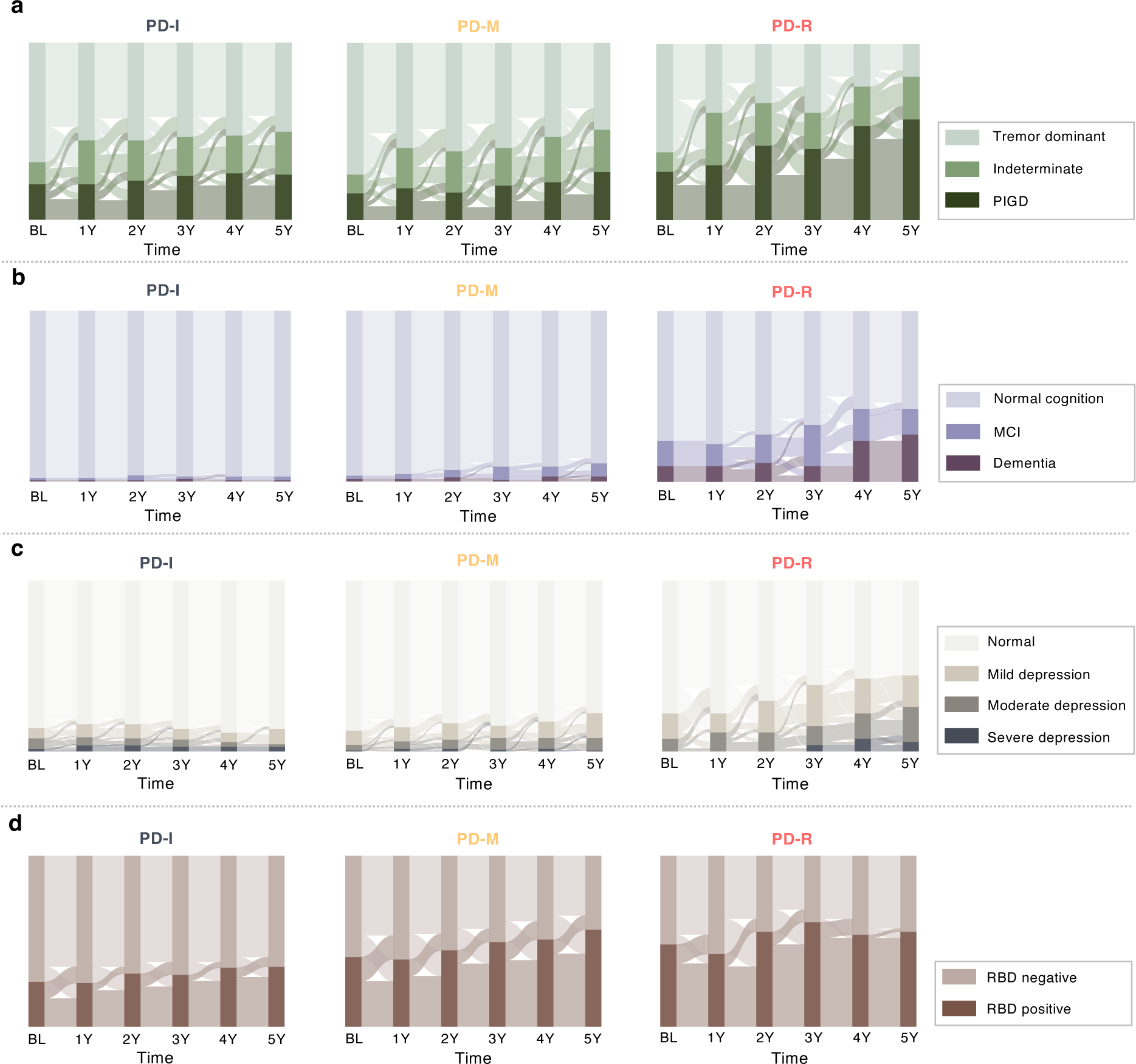
Sankey diagram showing evolution patterns of motor, cognition, mood, and sleep phenotypes by subtypes in the PPMI cohort. Abbreviations: MCI = mild cognition impairment; PD-I = Inching Pace PD subtype; PD-M = Moderate Pace PD subtype; PD-R = Rapid Pace PD subtype; PIGD = postural instability and gait disorder; RBD = REM (Rapid eye movement) sleep behaviour disorder.

**Extended Data Fig. 2.**
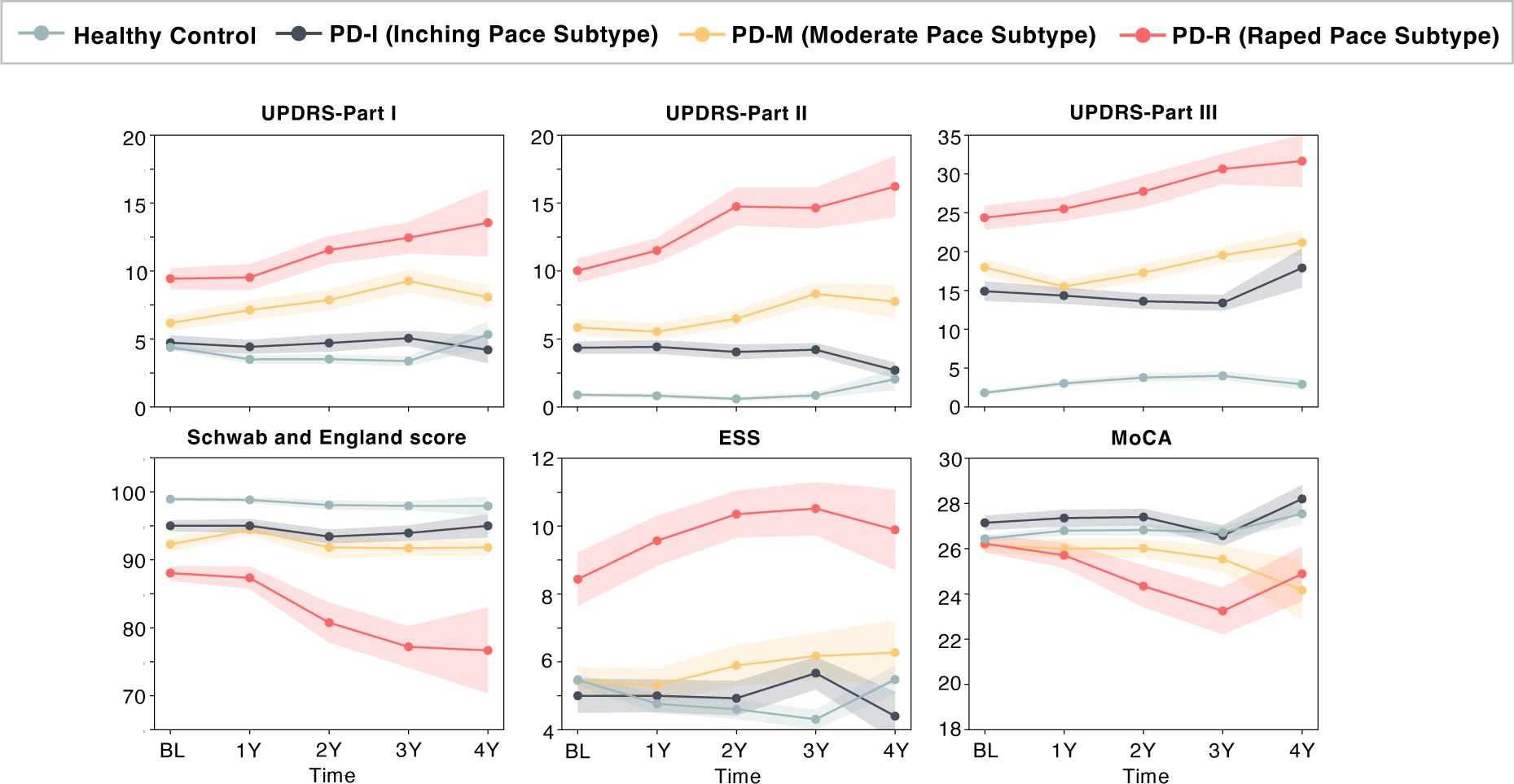
Averaged symptom progression trajectories by PD subtypes in the PDBP cohort. Abbreviations: ESS = Epworth sleepiness score; MDS-UPDRS = Movement Disorders Society–revised Unified Parkinson’s Disease Rating Scale; MoCA = Montreal Cognitive Assessment.

**Extended Data Fig. 3.**
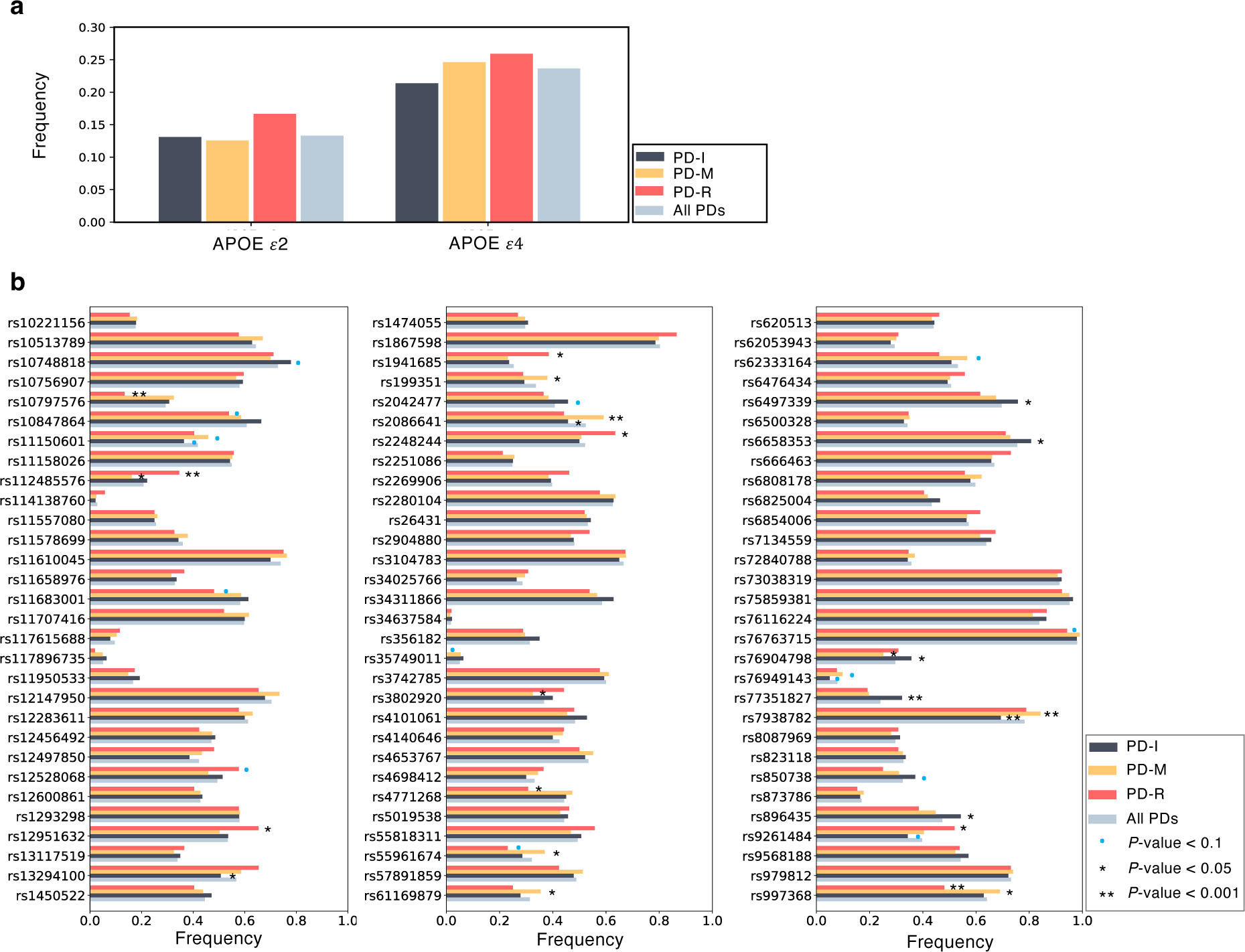
Results of genetic analysis across subtypes. Enrichment analysis didn’t find difference in APOE ε2 and ε4 alleles, GBA and LRRK2 variants among identified PD subtypes (**a**). Signals in 90 PD-related SNPs were found to be associated with the identified PD subtypes (**b**).

**Extended Data Fig. 4.**
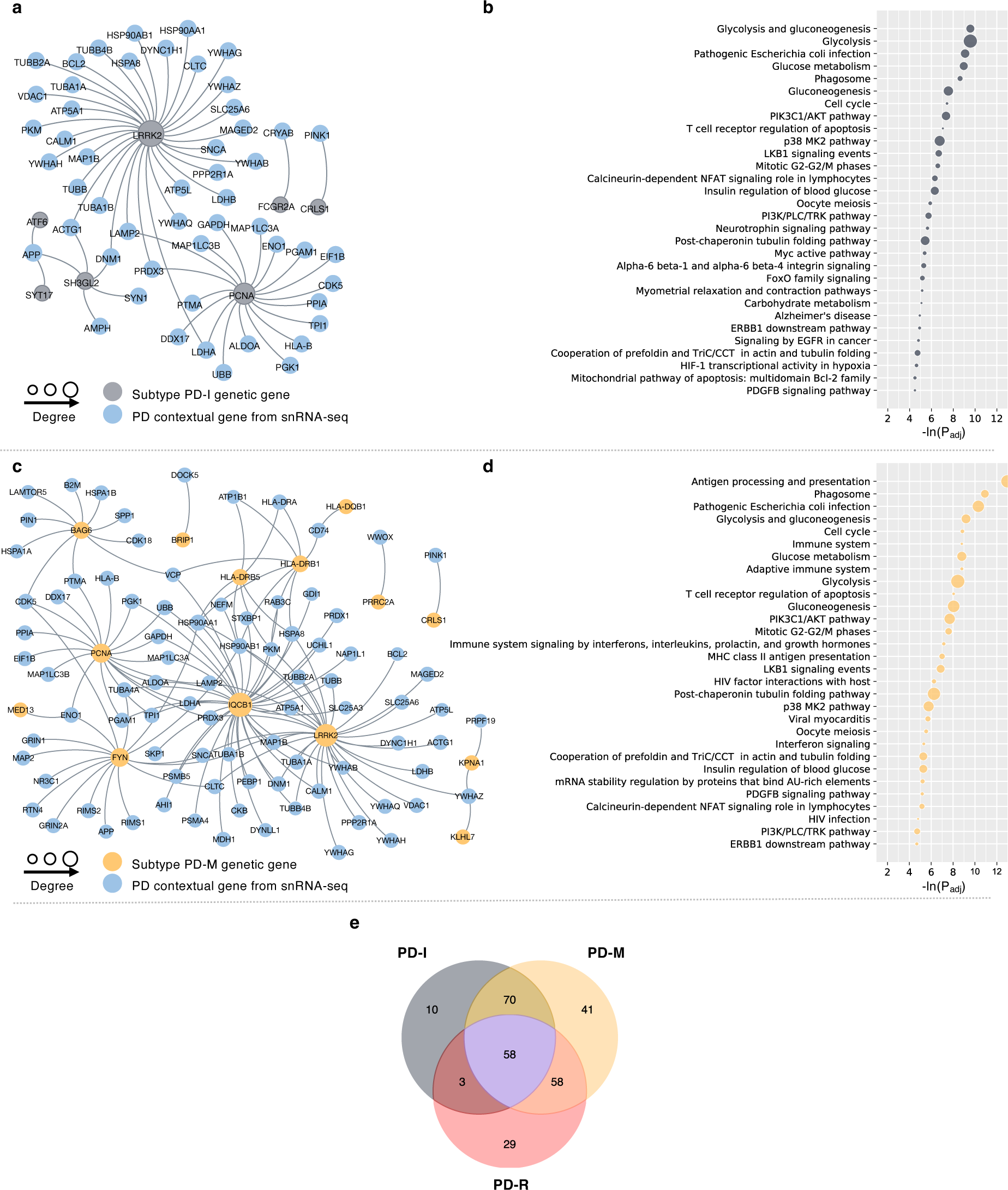
Genetic molecular modules of the subtypes. **a.** and **b.** Genetic molecular module and pathways enriched based on genetic molecular module of the PD-I subtype. **c.** and **d.** Genetic molecular module and pathways enriched based on genetic molecular module of PD-M. **e.** Venn plot showing overlaps of enriched pathways among the three subtypes.

**Extended Data Fig. 5.**
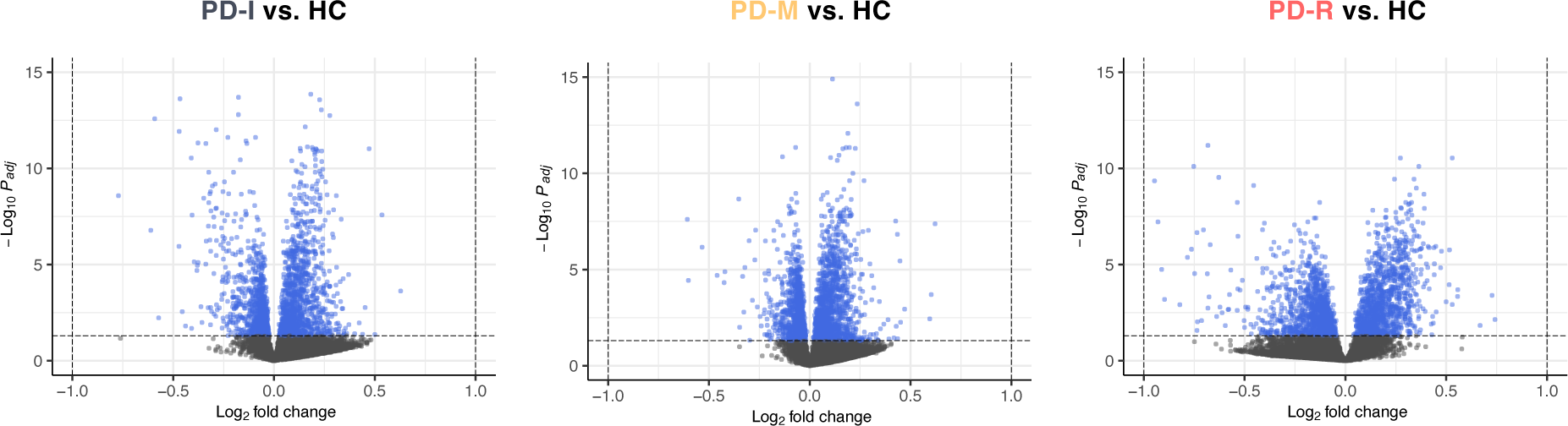
Volcano plots for differential gene expression analysis. Genes with adjusted *P* value (i.e., *Q* value) < 0.05 were considered as differentially expressed genes (DEGs) in each subtype (subtype vs. healthy controls [HCs]), which were further fed to the GPSnet algorithm for identifying gene modules of each of the identified PD subtypes.

**Extended Data Fig. 6.**
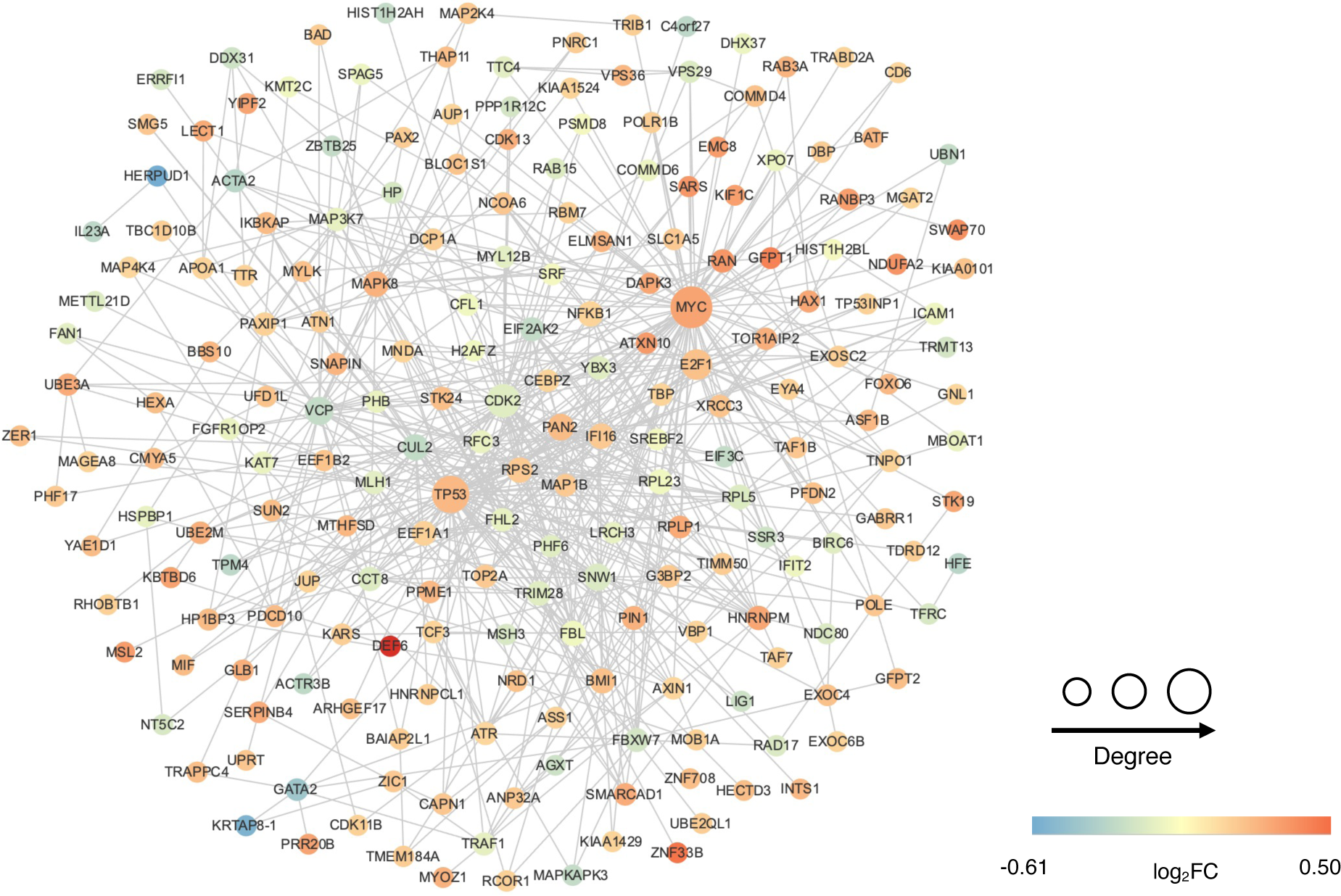
Transcriptomic molecular module of the PD-I subtype identified by gene expression analysis and network analysis (GPSnet) Color indicates log fold change in gene expression, PD-I vs. healthy control. Size of a gene indicates degrees (number of connected genes) of the gene in the protein-protein interaction network.

**Extended Data Fig. 7.**
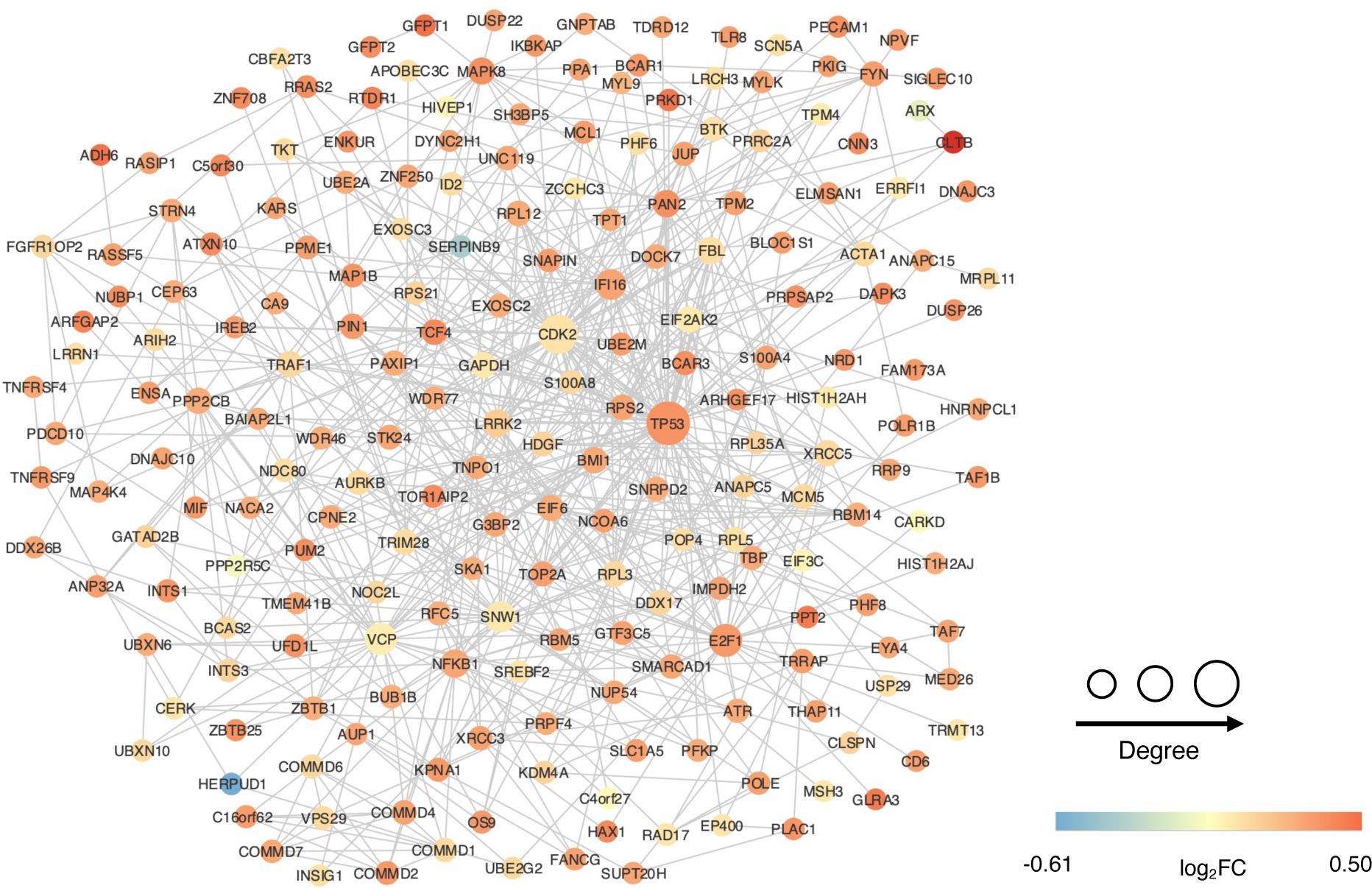
Transcriptomic molecular module of the PD-M subtype identified by gene expression analysis and network analysis (GPSnet) Color indicates log fold change in gene expression, PD-M vs. healthy control. Size of a gene indicates degrees (number of connected genes) of the gene in the protein-protein interaction network.

**Extended Data Fig. 8.**
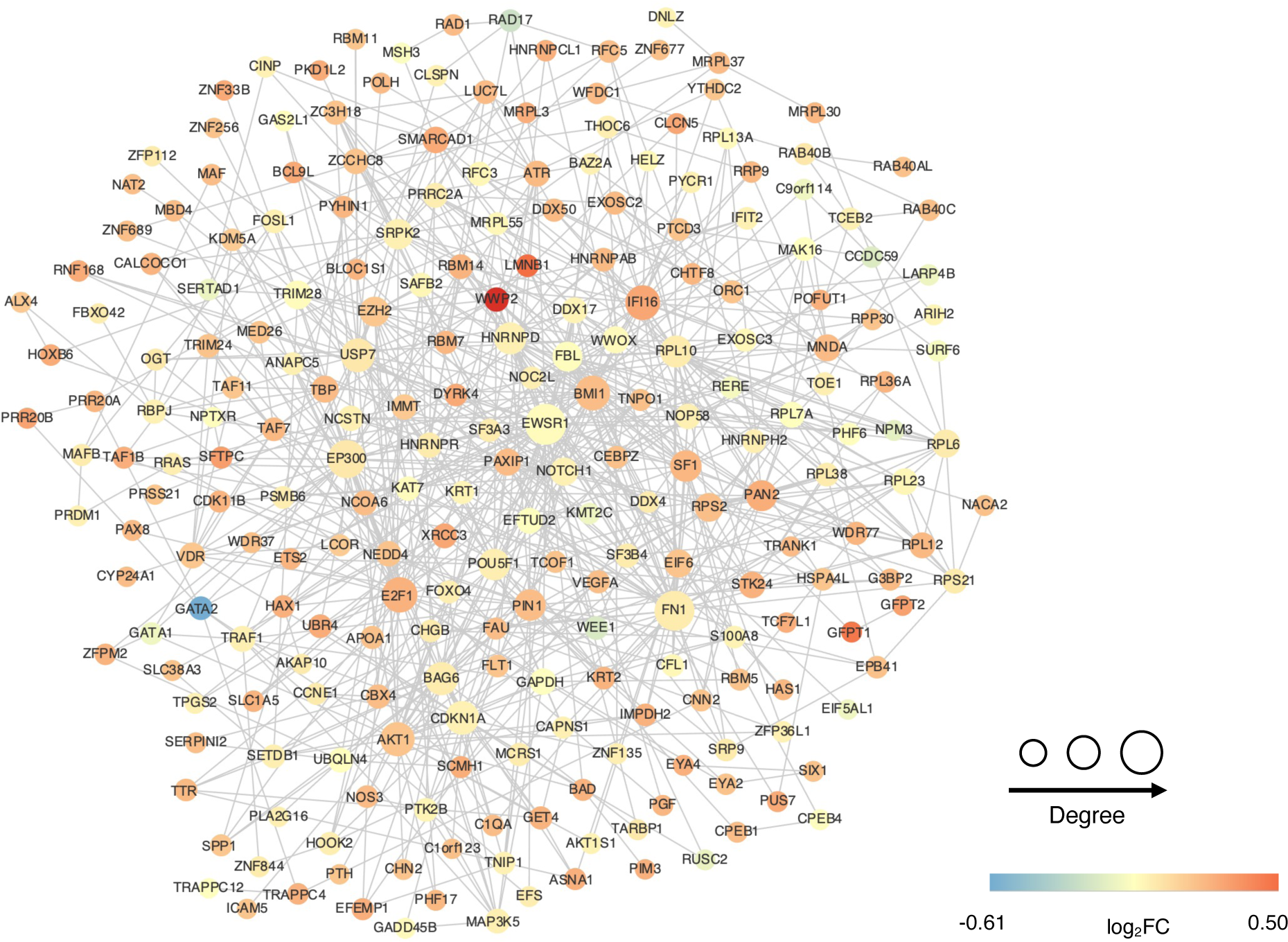
Transcriptomic molecular module of the PD-R subtype identified by gene expression analysis and network analysis (GPSnet) Color indicates log fold change in gene expression, PD-R vs. healthy control. Size of a gene indicates degrees (number of connected genes) of the gene in the protein-protein interaction network.

**Extended Data Fig. 9.**
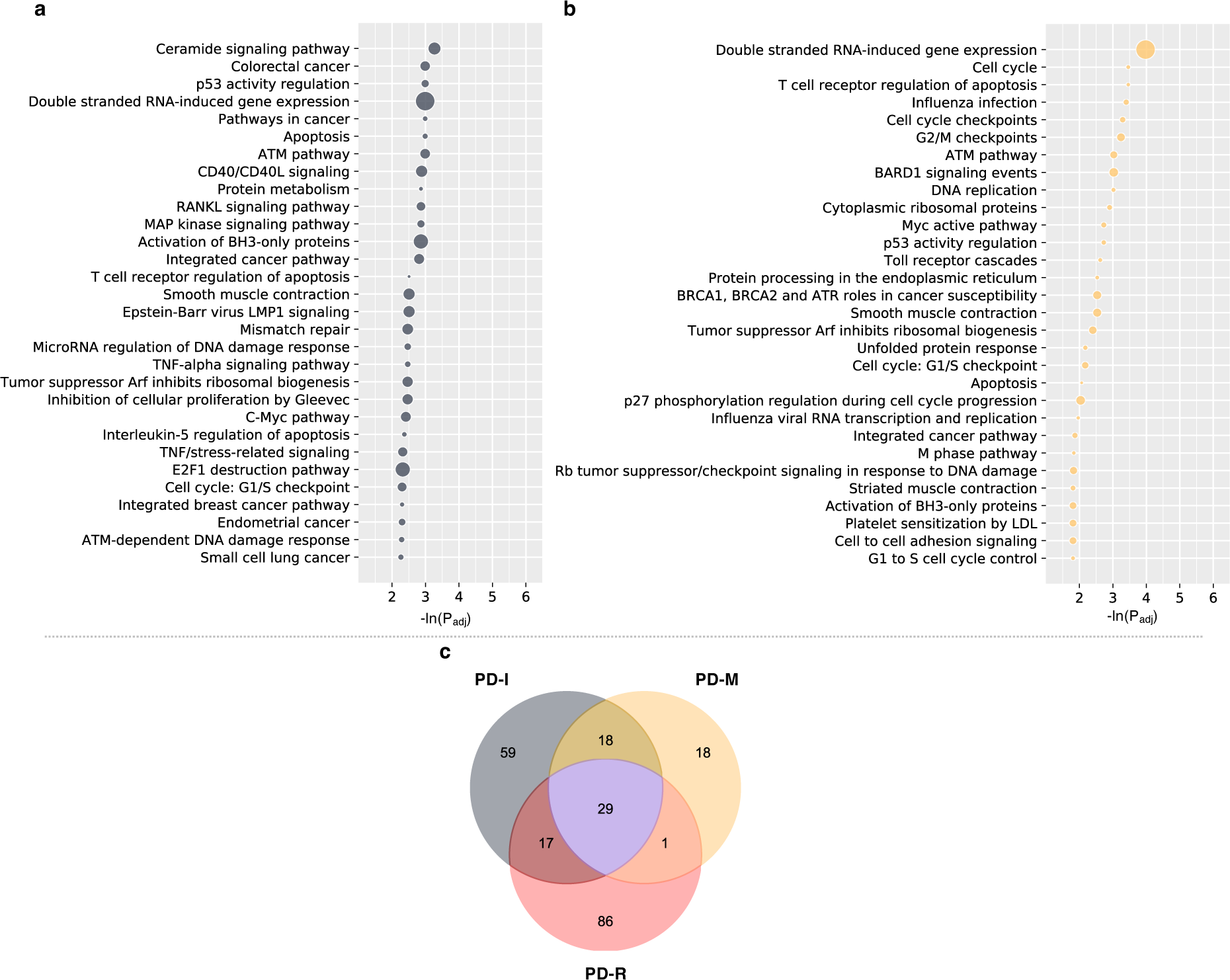
Enriched pathways based on transcriptomic molecular module of the subtypes. **a.** and **b.** Enriched pathways based on transcriptomic molecular module of the PD-I and PD-M subtypes. **c.** Venn plot showing overlaps of enriched pathways among the three subtypes.

**Extended Data Fig. 10.**
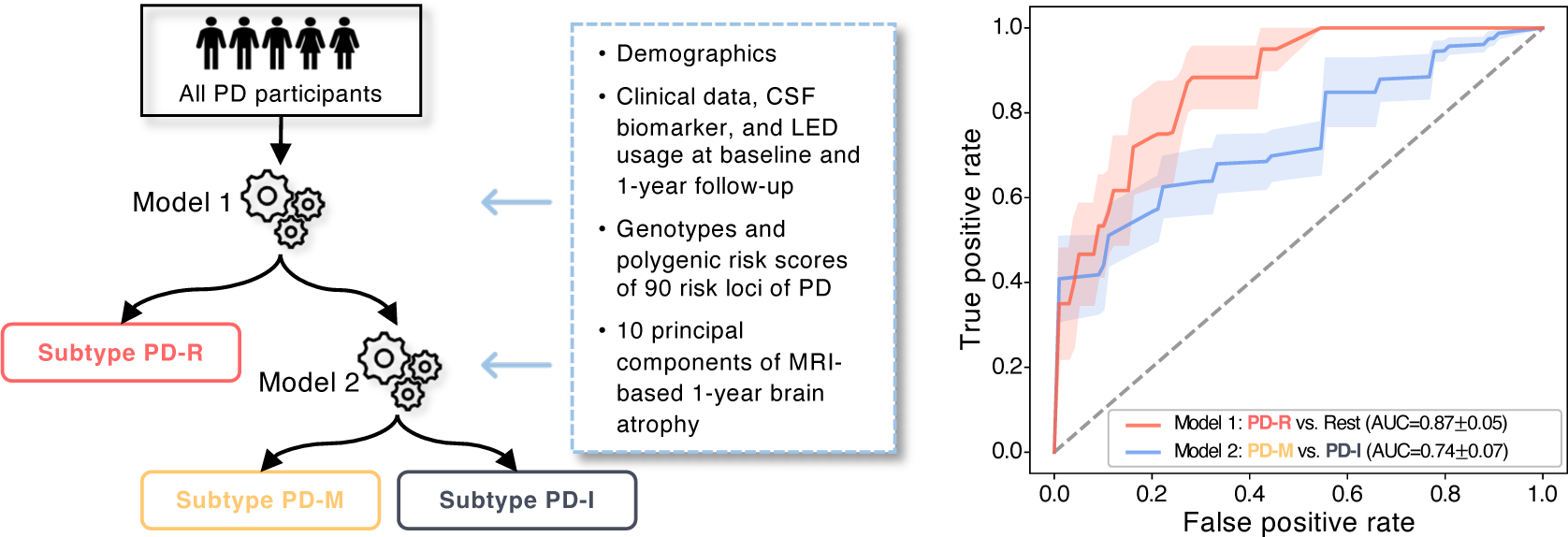
Classification model for separating the PD subtypes at early stage.

