## Supplemental file for "Identification of Parkinson PACE subtypes and repurposing treatments through integrative analyses of multimodal clinical progression, neuroimaging, genetic, and transcriptomic data"

#### Supplementary Methods

##### Bioinformatics analysis of single nucleus RNA-sequencing data

We utilized one set of human brain single nucleus RNA-sequencing data collected from 12 control donors with two brain regions: cortex and substantia nigra (SN) which included approximately 17,000 nuclei. It is available from Gene Expression Omnibus (https://www.ncbi.nlm.nih.gov/geo/) database with accession number GSE140231. We performed the bioinformatics analyses according to the processes described in the original manuscript[1]. Each brain region was analyzed individually, and the whole analyses were implemented with Seurat (4.0.6)[2]. Nuclei expressed with ≤ 500 genes, with ≥ 5% mitochondrial genes and ≥ 5% ribosomal genes were removed. Then the raw count was log-normalized and the top 2000 most variable genes were detected by function *FindVariableFeatures* with *selection.method* = '*vst*'. Next, all samples were integrated by functions *FindIntegrationAnchors* using canonical correlation analysis (CCA) and *InegrateData* with *dims = 1:42* and *1:25* for cortex and substantia nigra, separately. We then scaled the data and regressed out heterogeneity related with ribosomal, mitochondrial content, and number of UMIs. Clustering was performed with resolution 0.4 and 0.6 for cortex and substantia nigra, separately. We identified dopaminergic neuron with marker genes (*TH, LMX1B, KCNJ6, NR4A2* and *SLC6A3*) provided by the original manuscript[1] for substantia nigra only. DEGs for dopaminergic neuron were calculated against other cell types with MAST R package[3] regarding brain region substantia nigra.

### Supplementary Results – Determination of optimal cluster number

#### A. Subtype identification in the PPMI (development) cohort

Using learned representation vectors of participants in the PPMI cohort, dendrogram showed that the 3-cluster model is the optimal fit of the agglomerative hierarchical clustering model (see Supplementary Figure 1). In addition, out of 18 indices in 'NbClust', 8 suggested 3 clusters, 1 suggested 1 cluster, 4 suggested 2 clusters, 3suggested 4 clusters, and 2 suggested >7 clusters. In conclusion, by considering both dendrogram and the indices, the optimal cluster number was 3.

#### B. Subtype identification in the PDBP (validation) cohort

In the PDBP validation cohort, dendrogram also showed that the 3-cluster model is the optimal fit of the agglomerative hierarchical clustering model (see Supplementary Figure 2). In addition, out of 18 indices in 'NbClust', 5 suggested 3 clusters, 1 suggested 1 cluster, 5 suggested 2 clusters, and 5 suggested 4 clusters, and 1 suggested 8 clusters. In conclusion, by considering both dendrogram and the indices, the optimal cluster number was 3.

#### Supplementary Figures

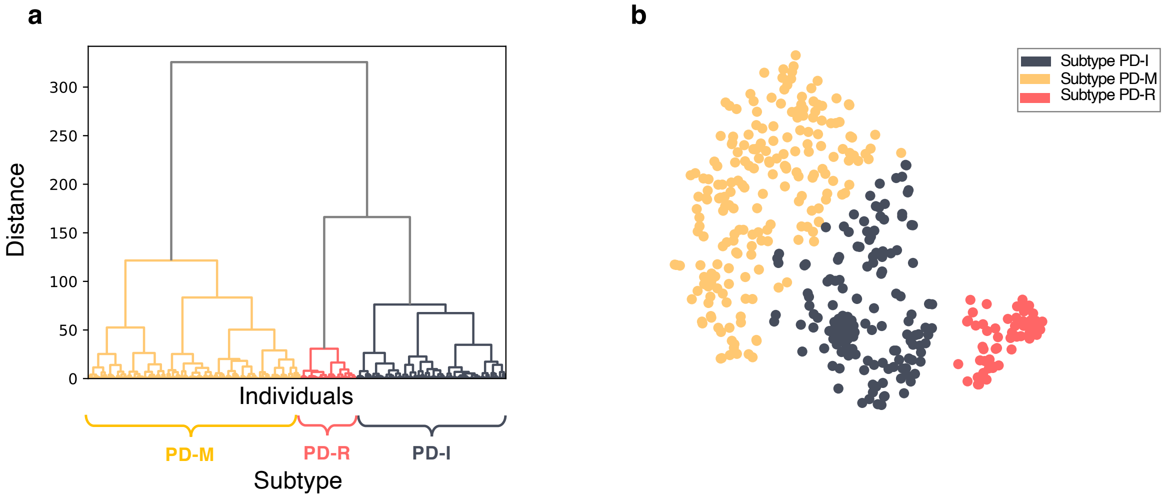

**Supplementary Fig. 1 Performance of hierarchical clustering in the PPMI cohort. a.** Dendrogram of hierarchical clustering shows clear three cluster structure of PDs in PPMI data. **b.** t-SNE visualization of shows clear three cluster structure of PDs in PPMI data.

Abbreviations: PD = Parkinson's disease; PD-I = Inching Pace PD subtype; PD-M = Moderate Pace PD subtype; PD-R = Rapid Pace PD subtype; PPMI = the Parkinson progression marker initiative; t-SNE = Student t-Distributed Stochastic Neighbor Embedding.

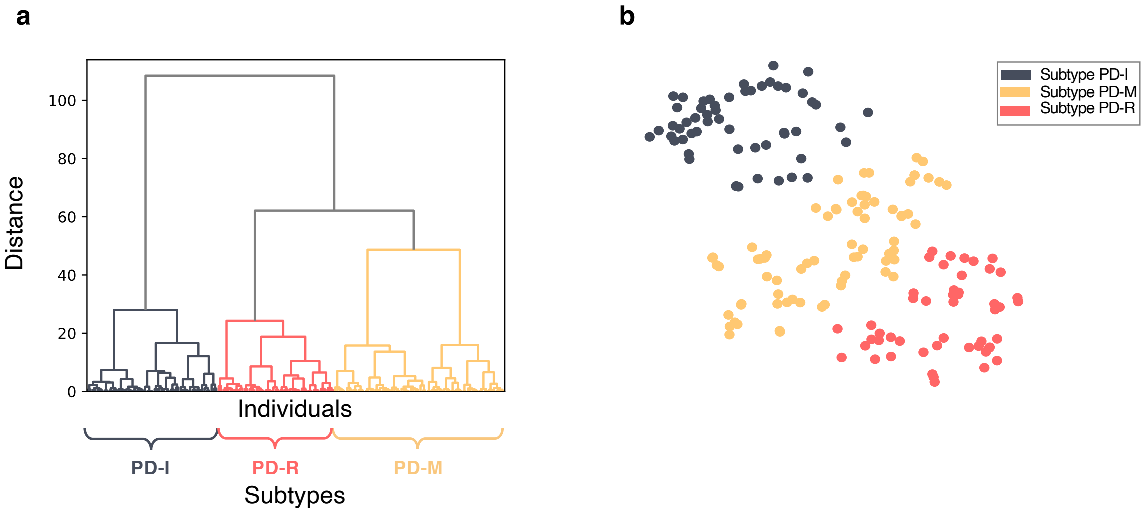

**Supplementary Fig. 2 Performance of hierarchical clustering in the PDBP cohort. a.** Dendrogram of hierarchical clustering shows clear three cluster structure of PDs in the PDBP data. **b.** t-SNE visualization of shows clear three cluster structure of PDs in the PDBP data.

Abbreviations: PD = Parkinson's disease; PDBP = the Parkinson's disease biomarkers program; PD-I = Inching Pace PD subtype; PD-M = Moderate Pace PD subtype; PD-R = Rapid Pace PD subtype; t-SNE = Student t-Distributed Stochastic Neighbor Embedding.

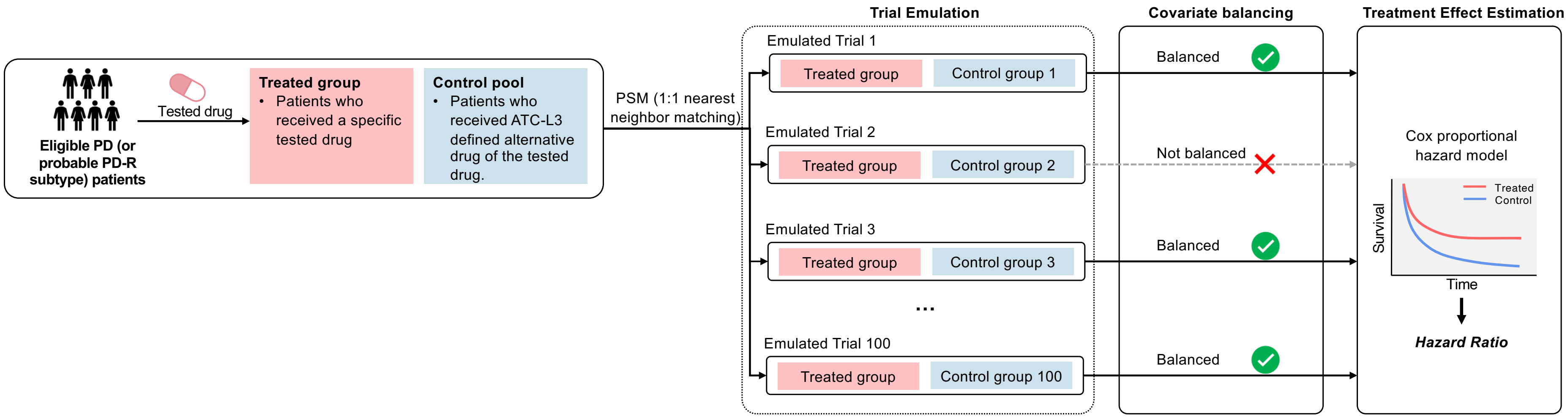

**Supplementary Fig. 3 Study pipeline of real-world patient data analysis for drug treatment effects estimation.**

Abbreviations: ATC-L3 = Anatomical Therapeutic Chemical level 3; PD = Parkinson's disease; PD-R Rapid Pace Parkinson's disease subtype.

### Supplementary Tables

**Supplementary Table 1. Characteristics and utilization of the studied cohorts**

| | Development cohort: PPMI | | | Validation cohort: PDBP | | |
|---|---|---|---|---|---|---|
| **Variables** | **HCs** | **SWEDD** | **PDs (de novo and treatment-naïve)** | **HCs** | **Early PDs** | **Other PDs** |
| **# of participants** | **188** | **61** | **406** | **211** | **210** | **287** |
| Demographics | | | | | | |
| Age at onset, year, mean (SD) | **-** | **-** | **59.6 (10.0)** | **-** | **63.0 (10.0)** | **57.6 (10.5)** |
| Sex male, N (%) | **121 (64.4)** | **38 (62.3)** | **266 (65.5)** | **100 (47.4)** | **120 (57.1)** | **182 (63.4)** |
| Race white, N (%) | **177 (94.1)** | **58 (95.1)** | **384 (94.6)** | **196 (92.9)** | **199 (94.8)** | **261 (90.9)** |
| Symptom duration, year (SD) | **-** | **-** | **0.6 (0.7)** | **-** | **1.0 (0.8)** | **8 (6.8)** |
| Family history (%) | **6 (3.2)** | **15 (24.6)** | **61 (15)** | **7 (3.3)** | **28 (13.3)** | **21 (7.3)** |
| Education history (%) | | | | | | |
| < 12 years | **5 (2.7)** | **10 (16.4)** | **26 (6.4)** | **2 (0.9)** | **7 (3.3)** | **4 (1.4)** |
| 12-16 years | **110 (58.5)** | **34 (55.7)** | **248 (61.1)** | **141 (66.8)** | **131 (62.4)** | **169 (58.9)** |
| ≥ 16 years | **73 (38.8)** | **17 (27.9)** | **132 (32.5)** | **68 (32.2)** | **71 (33.8)** | **113 (39.4)** |
| Utilization | | | | | | |
| Training deep progression embedding model | ✓ | ✓ | ✓ | ✓ | ✓ | ✓ |
| Clustering analysis for subtype identification | | | ✓ | | ✓ | |

Abbreviations: HC = Healthy controls; PD = Parkinson's disease; PDBP = Parkinson's Disease Biomarkers Program; PPMI = the Parkinson progression marker initiative; SD = standard deviation; SWEDD = subjects with scans without evidence for dopaminergic deficit.

**Supplementary Table 2. Clinical variables used for PD subtyping**

| Category | Data | Description | PPMI | PDBP |
|---|---|---|---|---|
| Motor assessment | MDS-UPDRS Part II[4] | Self-administered questionnaire of motor experiences of daily living. We used all items. | X | X |
| | MDS-UPDRS Part III[4] | Motor examination provided my rater. We used all items. | X | X |
| | Schwab-England activities of daily living score | Measure of the abilities of individuals living with PD relative to a completely independent situation. | X | X |
| Non-motor assessment | MDS-UPDRS Part I[4] | Non-motor experiences of daily living. We used all items. | X | X |
| | Scales for Outcomes in Parkinson's disease-Autonomic (SCOPA-AUT)[5] | The SCOPA-AUT was developed to evaluate autonomic symptoms. We used scores of the 7 domains, including gastrointestinal, urinary, cardiovascular, thermoregulatory, pupillomotor, and sexual. | X | |
| | Geriatric depression scale (GDS)[6] | Measure of depression in older adults. | X | |
| | Questionnaire for Impulsive-Compulsive Disorders in Parkinson's disease (QUIP)[7] | Measure of severity of symptoms and support a diagnosis of impulse control disorders and related disorders in PD. We used all items. | X | |
| | State-Trait Anxiety Inventory (STAI)[8] | The measure of trait and state anxiety. We used the STAI-Strait and STAI-State sub-scores. | X | |
| | Benton Judgment of Line Orientation (JOLO)[9] | A standardized measure of visuospatial judgment. We used the crude score and MOANS normative scores. | X | |
| | Hopkins Verbal Learning Test (HVLT)[10] | A memory test with six equivalent forms. | X | |
| | Letter-number sequencing (LNS) | A subset of Wechsler adult intelligence scale, measuring working memory, attention, mental control | X | |
| | Montreal Cognitive Assessment (MoCA)[11] | A screening assessment for detecting cognitive impairment. We used the visuospatial, naming, attention, language, delayed recall, abstraction, and verbal fluency sub-scores, and total MoCA score. | X | X |
| | Semantic verbal-language fluency test[12] | Assessment of semantic knowledge, retrieval ability, and executive functioning. We used the sub-scores in terms of animals, vegetables, and fruits. | X | |
| | Symbol–Digit Matching (SDM)[13] | A neuropsychological test that examines a person's attention and speed of processing. | X | |
| | Epworth Sleepiness Score (ESS)[14] | Measure of daytime sleepiness. We used all items. | X | X |
| | REM sleep behaviour disorder (RBD)[15] | A questionnaire for RBD. We used all items. | X | X |
| | Cranial Nerve Examination | A kind of neurological examination that is used to identify problems with the cranial nerves. We used the 9 components. | X | |
| PD medication | Levodopa equivalent daily dose | Levodopa equivalent daily dose | X | |

Abbreviations: MDS-UPDRS = Movement Disorders Society–revised Unified Parkinson's Disease Rating Scale.

**Supplementary Table 3. 2-year follow-up clinical characteristics by subtypes in the PPMI cohort**

| Variables | Subtype PD-I (Inching Pace) | Subtype PD-M (Moderate Pace) | Subtype PD-R (Rapid Pace) | P-value[a] | Post-hoc[b] | P-value adjusted[c] |
|---|---|---|---|---|---|---|
| **# of participants** | **145** | **207** | **54** | **-** | **-** | **-** |
| ***Motor manifestations*** | | | | | | |
| MDS-UPDRS Part II, mean (SD) | 6.0 (4.6) | 8.2 (4.9) | 12.1 (6.0) | <0.001 | All comparisons | <0.001 |
| MDS-UPDRS Part III, mean (SD) | 23.9 (10.6) | 27.1 (10.5) | 33.4 (13.1) | <0.001 | All comparisons | <0.001 |
| H&Y Stage, mean (SD) | 1.7 (0.6) | 1.8 (0.5) | 2.1 (0.6) | <0.001 | III vs. rest | 0.028 |
| Schwab and England score, mean (SD) | 90.6 (7.2) | 89.0 (7.5) | 83.5 (9.7) | <0.001 | III vs. rest | <0.001 |
| Tremor score, mean (SD) | 0.5 (0.4) | 0.6 (0.4) | 0.6 (0.5) | 0.006 | I vs. II | 0.030 |
| PIGD score, mean (SD) | 0.3 (0.3) | 0.3 (0.3) | 0.7 (0.6) | <0.001 | III vs. rest | <0.001 |
| Motor phenotype, N (%) | | | | | | |
| Tremor | 73 (50.3) | 127 (61.4) | 18 (33.3) | <0.001 | - | - |
| Indeterminate | 30 (20.7) | 48 (23.2) | 13 (24.1) | | | |
| PIGD | 29 (20.0) | 32 (15.5) | 23 (42.6) | | | |
| ***Non-motor manifestations*** | | | | | | |
| MDS-UPDRS Part I, mean (SD) | 6.8 (4.3) | 7.4 (5.1) | 10.7 (5.4) | <0.001 | III vs. rest | <0.001 |
| Hallucination, mean (SD) | 0.09 (0.32) | 0.05 (0.21) | 0.24 (0.62) | 0.002 | III vs. rest | 0.001 |
| Apathy, mean (SD) | 0.3 (0.7) | 0.4 (0.7) | 0.6 (0.9) | 0.088 | - | 0.205 |
| Pain, mean (SD) | 0.8 (0.8) | 0.9 (0.9) | 1.1 (1.0) | 0.076 | - | 0.016 |
| Fatigue, mean (SD) | 0.6 (0.7) | 0.8 (0.9) | 1.2 (1.1) | <0.001 | III vs. rest | <0.001 |
| Sleep, mean (SD) | | | | | | |
| Epworth sleepiness score | 5.1 (3.4) | 7.3 (4.1) | 8.5 (4.8) | <0.001 | I vs. rest | <0.001 |
| REM sleep behavior disorder | 3.9 (2.8) | 4.7 (3.0) | 5.4 (3.4) | 0.006 | 1 vs. III | 0.007 |
| Sleep phenotype, missing = 1, N (%) | | | | | | |
| REM sleep behavior disorder positive | 45 (31.0) | 92 (44.7) | 30 (55.6) | 0.002 | - | - |
| REM sleep behavior disorder negative | 100 (69.0) | 114 (55.3) | 24 (44.4) | | | |
| QUIP (Impulse control disorders) , mean (SD) | 0.3 (0.7) | 0.3 (0.7) | 0.3 (0.8) | 0.830 | - | 0.929 |
| Geriatric depression scale, mean (SD) | 2.3 (2.8) | 2.7 (3.0) | 3.4 (2.6) | 0.064 | - | 0.052 |
| Depression phenotype, missing = 1, N (%) | | | | | | |
| Normal | 122 (84.1) | 172 (83.5) | 38 (70.4) | 0.121 | - | - |
| Mild | 12 (8.3) | 20 (9.7) | 10 (18.5) | | | |
| Moderate | 6 (4.1) | 11 (5.3) | 6 (11.1) | | | |
| Severe | 5 (3.5) | 3 (1.5) | 0 (0) | | | |

| | | | | | | |
|---|---|---|---|---|---|---|
| State trait anxiety index, mean (SD) | | | | | | |
| State subscore | 31.7 (10.4) | 32.2 (9.3) | 36.3 (11.6) | 0.016 | III vs. rest | 0.007 |
| Trait subscore | 31.7 (10.3) | 32.5 (8.8) | 35.3 (10.3) | 0.075 | - | 0.012 |
| SCOPA autonomic questionnaire, mean (SD) | | | | | | |
| Gastrointestinal (up+down) | 2.6 (2.3) | 2.8 (2.3) | 4.3 (2.6) | <0.001 | III vs. rest | <0.001 |
| Urinary | 4.0 (2.5) | 4.7 (2.9) | 6.5 (7.6) | <0.001 | III vs. rest | 0.016 |
| Cardiovascular | 0.6 (0.8) | 0.7 (1.1) | 0.9 (1.2) | 0.083 | - | 0.128 |
| Thermoregulatory | 0.4 (0.8) | 0.6 (1.0) | 0.4 (0.7) | 0.280 | - | 0.208 |
| Pupillomotor | 0.4 (0.7) | 0.5 (0.7) | 0.6 (0.8) | 0.532 | - | 0.320 |
| Skin | 0.8 (1.1) | 0.9 (1.0) | 0.9 (1.1) | 0.727 | - | 0.312 |
| Sexual | 5.0 (6.8) | 4.3 (6.0) | 5.8 (6.7) | 0.274 | - | 0.559 |
| Total (sum all) | 13.9 (8.8) | 14.4 (9.2) | 19.4 (11.1) | 0.001 | III vs. rest | <0.001 |
| Cognitive function, mean (SD) | | | | | | |
| MoCA-visuospatial | 4.5 (0.8) | 4.3 (0.8) | 3.7 (1.5) | <0.001 | III vs. rest | <0.001 |
| MoCA-naming | 2.9 (0.2) | 2.9 (0.3) | 2.9 (0.4) | 0.244 | - | 0.363 |
| MoCA-attention | 5.8 (0.5) | 5.6 (0.7) | 5.3 (1.0) | <0.001 | III vs. rest | <0.001 |
| MoCA-language | 2.6 (0.5) | 2.4 (0.8) | 2.0 (1.0) | <0.001 | All comparisons | <0.001 |
| MoCA-delayed recall | 3.7 (1.5) | 3.0 (1.7) | 2.0 (1.9) | <0.001 | All comparisons | <0.001 |
| MoCA total score | 27.7 (2.4) | 26.2 (2.7) | 23.6 (4.4) | <0.001 | All comparisons | <0.001 |
| Benton judgment of line orientation | 13.3 (1.8) | 12.8 (2.1) | 11.4 (3.1) | <0.001 | III vs. rest | <0.001 |
| HVLT-total recall | 25.6 (4.7) | 23.5 (5.3) | 19.7 (6.1) | <0.001 | All comparisons | <0.001 |
| HVLT-delayed recall | 8.9 (2.5) | 8.3 (2.9) | 6.1 (3.4) | <0.001 | III vs. rest | <0.001 |
| HVLT-discrimination recognition | 11.0 (2.3) | 10.7 (2.2) | 9.7 (3.1) | 0.005 | III vs. rest | 0.047 |
| HVLT-retention | 0.9 (0.2) | 0.9 (0.2) | 0.7 (0.3) | <0.001 | III vs. rest | 0.010 |
| LNS | 11.2 (2.6) | 10.2 (2.5) | 8.4 (3.4) | <0.001 | All comparisons | <0.001 |
| Semantic fluency | 53.7 (13.4) | 48.3 (11.9) | 39.4 (11.2) | <0.001 | All comparisons | <0.001 |
| Symbol digit test | 48 (9.6) | 43.9 (9.7) | 38.0 (10.8) | <0.001 | All comparisons | <0.001 |
| Cognitive phenotype, missing = 7, N (%) | | | | | | |
| Normal | 133 (96.4) | 193 (93.2) | 39 (72.2) | <0.001 | - | - |
| MCI | 4 (2.8) | 9 (4.4) | 9 (16.7) | | | |
| Dementia | 1 (0.7) | 5 (2.4) | 6 (11.1) | | | |

[a] P-values were calculated using ANOVA (for continuous variables) and $\chi^2$ test (for categorical variables) where appropriate.
[b] Post-hoc analysis was performed using the Tukey HSD test when the ANOVA P-value < 0.05.
[c] ANCOVA was used to calculate p-values (for continuous variables) adjusting for age and sex.

Abbreviations: HVLT = Hopkins Verbal Learning Test; MCI = mild cognitive impairment; MDS-UPDRS = Movement Disorders Society–revised Unified Parkinson's Disease Rating Scale; MoCA = Montreal Cognitive Assessment; PIGD = postural instability and gait disorder; PPMI = the Parkinson's Progression Markers Initiative; SCOPA = Scales for Outcomes in Parkinson's Disease.

**Supplementary Table 4. 5-year follow-up clinical characteristics by subtypes in the PPMI cohort**

| **Variables** | **Subtype PD-I (Inching Pace)** | **Subtype PD-M (Moderate Pace)** | **Subtype PD-R (Rapid Pace)** | **P-value[a]** | **Post-hoc[b]** | **P-value adjusted[c]** |
|---|---|---|---|---|---|---|
| **# of participants** | **145** | **207** | **54** | **-** | **-** | **-** |
| ***Motor manifestations*** | | | | | | |
| MDS-UPDRS Part II, mean (SD) | 5.7 (4.3) | 11.1 (5.6) | 16.5 (10.0) | <0.001 | All comparisons | <0.001 |
| MDS-UPDRS Part III, mean (SD) | 23.0 (9.5) | 32.5 (12.9) | 38.9 (15.8) | <0.001 | All comparisons | <0.001 |
| H&Y Stage, mean (SD) | 1.9 (0.4) | 2.0 (0.4) | 2.5 (0.9) | <0.001 | III vs. rest | <0.001 |
| Schwab and England score, mean (SD) | 90.5 (6.6) | 84.1 (9.3) | 70.9 (23.6) | <0.001 | All comparisons | <0.001 |
| Tremor score, mean (SD) | 0.5 (0.4) | 0.7 (0.5) | 0.6 (0.5) | <0.001 | I vs. II | 0.003 |
| PIGD score, mean (SD) | 0.3 (0.3) | 0.5 (0.4) | 1.2 (1.0) | <0.001 | All comparisons | <0.001 |
| Motor phenotype, N (%) | | | | | | |
| Tremor | 54 (37.2) | 102 (49.3) | 8 (14.8) | 0.004 | - | - |
| Indeterminate | 24 (16.6) | 49 (23. 7) | 6 (11.1) | | | |
| PIGD | 21(14.5) | 56 (27.1) | 20 (37.0) | | | |
| ***Non-motor manifestations*** | | | | | | |
| MDS-UPDRS Part I, mean (SD) | 6.5 (4.7) | 9.8 (5.6) | 14.3 (9.0) | <0.001 | All comparisons | <0.001 |
| Hallucination, mean (SD) | 0.1 (0.3) | 0.2 (0.4) | 0.4 (1.1) | 0.002 | III vs. rest | 0.005 |
| Apathy, mean (SD) | 0.2 (0.5) | 0.5 (0.8) | 0.9 (1.2) | <0.001 | All comparisons | <0.001 |
| Pain, mean (SD) | 0.6 (0.9) | 1.0 (1.1) | 1.4 (1.1) | 0.002 | I vs. rest | <0.001 |
| Fatigue, mean (SD) | 0.7 (0.8) | 1.1 (1.0) | 1.6 (1.2) | <0.001 | All comparisons | <0.001 |
| Sleep, mean (SD) | | | | | | |
| Epworth sleepiness score | 5.4 (3.9) | 8.5 (4.4) | 10.2 (5.7) | <0.001 | I vs. rest | <0.001 |
| REM sleep behavior disorder | 3.7 (2.9) | 5.3 (3.1) | 5.7 (3.6) | <0.001 | I vs. rest | <0.001 |
| Sleep phenotype, missing = 1, N (%) | | | | | | |
| REM sleep behavior disorder positive | 51 (35.2) | 117 (56.8) | 30 (55.6) | <0.001 | - | - |
| REM sleep behavior disorder negative | 94 (64.8) | 89 (43.2) | 24 (44.4) | | | |
| QUIP (Impulse control disorders) , mean (SD) | 0.3 (0.7) | 0.5 (0.9) | 0.2 (0.4) | 0.028 | | 0.054 |
| Geriatric depression scale, mean (SD) | 1.6 (1.9) | 3.0 (2.6) | 4.9 (4.2) | <0.001 | All comparisons | <0.001 |
| Depression phenotype, missing = 1, N (%) | | | | | | |
| Normal | 126 (86.9) | 160 (77.7) | 30 (55.6) | <0.001 | - | - |
| Mild | 13 (9.0) | 30 (14.6) | 10 (18.5) | | | |
| Moderate | 2 (1.4) | 15 (7.3) | 11 (20.4) | | | |

| | | | | | | |
|---|---|---|---|---|---|---|
| Severe | 4 (2.7) | 1 (0.5) | 3 (5.6) | | | |
| State trait anxiety index, mean (SD) | | | | | | |
| State subscore | 28.8 (8.9) | 33.0 (9.5) | 36.1 (13.4) | <0.001 | I vs. rest | <0.001 |
| Trait subscore | 29.5 (9.2) | 33.7 (9.7) | 36.8 (13.5) | <0.001 | I vs. rest | <0.001 |
| SCOPA autonomic questionnaire, mean (SD) | | | | | | |
| Gastrointestinal (up+down) | 2.7 (2.2) | 3.8 (2.5) | 5.1 (3.7) | <0.001 | All comparisons | <0.001 |
| Urinary | 4.4 (2.7) | 5.8 (5.3) | 6.7 (4.8) | 0.017 | I vs. rest | 0.111 |
| Cardiovascular | 0.5 (0.9) | 0.8 (1.2) | 1.4 (1.8) | 0.001 | III vs. rest | 0.002 |
| Thermoregulatory | 0.5 (1.1) | 0.8 (1.2) | 0.4 (0.7) | 0.054 | - | 0.032 |
| Pupillomotor | 0.4 (0.7) | 0.6 (0.8) | 0.8 (1.0) | 0.048 | - | 0.062 |
| Skin | 0.7 (1.0) | 1.2 (1.2) | 1.5 (1.9) | <0.001 | I vs. rest | <0.001 |
| Sexual | 4.5 (6.3) | 5.2 (6.4) | 6.7 (7.4) | 0.248 | - | 0.085 |
| Total (sum all) | 13.7 (8.7) | 18.3 (11.1) | 22.5 (15.2) | <0.001 | I vs. rest | <0.001 |
| Cognitive function, mean (SD) | | | | | | |
| MoCA-visuospatial | 4.6 (0.7) | 4.2 (1.1) | 3.6 (1.6) | <0.001 | All comparisons | <0.001 |
| MoCA-naming | 3 (0.2) | 2.9 (0.2) | 2.8 (0.6) | 0.041 | III vs. rest | 0.054 |
| MoCA-attention | 5.7 (0.5) | 5.5 (0.8) | 4.8 (1.4) | <0.001 | III vs. rest | <0.001 |
| MoCA-language | 2.6 (0.6) | 2.5 (0.7) | 2.2 (0.9) | 0.013 | I vs. III | 0.028 |
| MoCA-delayed recall | 4.3 (1.1) | 3.3 (1.6) | 1.9 (1.8) | <0.001 | All comparisons | <0.001 |
| MoCA total score | 28.3 (1.7) | 26.5 (3.2) | 23.0 (5.5) | <0.001 | All comparisons | <0.001 |
| Benton judgment of line orientation | 13.0 (2) | 12.3 (2.2) | 11.0 (2.8) | <0.001 | All comparisons | <0.001 |
| HVLT-total recall | 27.6 (5.1) | 24.1 (6.3) | 17.2 (4.9) | <0.001 | All comparisons | <0.001 |
| HVLT-delayed recall | 10 (2.4) | 8.4 (3.1) | 4.9 (3.1) | <0.001 | All comparisons | <0.001 |
| HVLT-discrimination recognition | 11.3 (1.1) | 10.5 (2.0) | 8.9 (2.5) | <0.001 | All comparisons | <0.001 |
| HVLT-retention | 0.9 (0.2) | 0.9 (0.2) | 0.7 (0.4) | <0.001 | All comparisons | <0.001 |
| LNS | 11.5 (2.6) | 9.7 (2.8) | 8.0 (3.4) | <0.001 | All comparisons | <0.001 |
| Semantic fluency | 55.6 (12.4) | 47.1 (11.5) | 35.3 (13.4) | <0.001 | All comparisons | <0.001 |
| Symbol digit test | 50.4 (10.7) | 44.1 (10.7) | 39.4 (15.3) | <0.001 | I vs. rest | <0.001 |
| Cognitive phenotype, missing = 7, N (%) | | | | | | |
| Normal | 134 (97.1) | 185 (89.4) | 31 (51.4) | | | |
| MCI | 3 (2.2) | 16 (7.7) | 8 (14.8) | <0.001 | - | - |
| Dementia | 1 (0.7) | 6 (2.9) | 15 (27.8) | | | |

[a] P-values were calculated using ANOVA (for continuous variables) and $\chi^2$ test (for categorical variables) where appropriate.
[b] Post-hoc analysis was performed using the Tukey HSD test when the ANOVA P-value < 0.05.
[c] ANCOVA was used to calculate p-values (for continuous variables) adjusting for age, sex, and levodopa equivalent daily dose.

Abbreviations: HVLT = Hopkins Verbal Learning Test; MCI = mild cognitive impairment; MDS-UPDRS = Movement Disorders Society–revised Unified Parkinson's Disease Rating Scale; MoCA = Montreal Cognitive Assessment; PIGD = postural instability and gait disorder; PPMI = the Parkinson's Progression Markers Initiative; SCOPA = Scales for Outcomes in Parkinson's Disease.

**Supplementary Table 5. Demographics and baseline clinical characteristics by subtypes in the PDBP cohort**

| Variables | Subtype PD-I (Inching Pace) | Subtype PD-M (Moderate Pace) | Subtype PD-R (Rapid Pace) | P-value[a] | Post-hoc[b] | P-value adjusted[c] |
|---|---|---|---|---|---|---|
| **# of participants** | **55** | **72** | **49** | **-** | **-** | **-** |
| Age at onset, year, mean (SD) | 58.6 (9.8) | 64.8 (9.3) | 65.2 (9.8) | <0.001 | I vs. rest | - |
| Sex male, N (%) | 28 (50.9) | 43 (59.7) | 32 (65.3) | 0.335 | - | - |
| Race white, N (%) | 54 (98.2) | 66 (91.7) | 47 (95.9) | 0.232 | - | - |
| Symptom duration, year, mean (SD) | 0.9 (0.8) | 0.8 (0.7) | 1.1 (0.8) | 0.183 | - | - |
| Family history, N (%) | 7 (12.7) | 10 (13.4) | 8 (16.3) | 0.896 | - | - |
| Education history, N (%) | | | | | | |
| Less than 12 years | - | 2 (2.8) | 1 (2.0) | 0.257 | - | - |
| 12-16 years | 32 (58.2) | 50 (69.4) | 27 (55.1) | | | |
| Greater than 16 years | 23 (41.8) | 20 (27.8) | 20 (40.8) | | | |
| ***Motor manifestations*** | | | | | | |
| MDS-UPDRS Part II, mean (SD) | 3.9 (2.9) | 5.6 (4.7) | 9.5 (6.0) | <0.001 | III vs. rest | <0.001 |
| MDS-UPDRS Part III, mean (SD) | 14.9 (8.9) | 18.1 (8.9) | 24.4 (10.4) | <0.001 | III vs. rest | 0.012 |
| H&Y Stage, mean (SD) | 1.7 (0.5) | 1.8 (0.5) | 2.1 (0.5) | <0.001 | III vs. rest | <0.001 |
| Schwab and England score, mean (SD) | 94.4 (5.0) | 91.9 (7.8) | 88.2 (6.7) | <0.001 | III vs. rest | <0.001 |
| Tremor score, mean (SD) | 0.4 (0.4) | 0.5 (0.4) | 0.4 (0.3) | 0.169 | - | 0.186 |
| PIGD score, mean (SD) | 0.2 (0.2) | 0.3 (0.2) | 0.6 (0.5) | <0.001 | III vs. rest | <0.001 |
| Motor phenotype, N (%) | | | | | | |
| Tremor | 37 (67.3) | 48 (66.7) | 18 (36.7) | 0.002 | III vs. rest | - |
| Indeterminate | 4 (7.2) | 8 (11.1) | 4 (8.2) | | | |
| PIGD | 14 (25.5) | 16 (22.2) | 27 (55.1) | | | |
| ***Non-motor manifestations*** | | | | | | |
| MDS-UPDRS Part I, mean (SD) | 4.7 (3.3) | 6.2 (4.4) | 9.1 (5.0) | <0.001 | III vs. rest | <0.001 |
| Hallucination, mean (SD) | 0.02 (0.12) | 0.03 (0.17) | 0.14 (0.35) | 0.009 | III vs. rest | 0.010 |
| Apathy, mean (SD) | 0.2 (0.4) | 0.2 (0.5) | 0.4 (0.8) | 0.201 | - | 0.170 |
| Pain, mean (SD) | 0. 8 (0.9) | 0.7 (0.7) | 1.0 (1.1) | 0.116 | - | 0.076 |
| Fatigue, mean (SD) | 0.7 (0.6) | 0.8 (0.8) | 1.1 (0.9) | 0.039 | I vs. III | 0.037 |
| Sleep, mean (SD) | | | | | | |
| Epworth sleepiness score | 5.1 (3.2) | 5.4 (3.2) | 8.3 (5.1) | <0.001 | I vs. III | <0.001 |
| REM sleep behavior disorder | 0.1 (0.8) | 0.08 (0.4) | 0.1 (0.7) | 0.885 | - | 0.817 |
| Cognitive function, mean (SD) | | | | | | |

| | | | | | | |
|---|---|---|---|---|---|---|
| MoCA-language | 2.5 (0.8) | 2.4 (0.7) | 2.3 (0.8) | 0.401 | - | 0.575 |
| MoCA total score | 27.1 (2.2) | 26.2 (2.4) | 26.2 (2.5) | 0.050 | - | 0.361 |

[a] P-values were calculated using ANOVA (for continuous variables) and $\chi^2$ test (for categorical variables) where appropriate.
[b] Post-hoc analysis was performed using the Tukey HSD test when the ANOVA P-value < 0.05.
[c] ANCOVA was used to calculate p-values (for continuous variables) adjusting for age and sex.

Abbreviations: MDS-UPDRS = Movement Disorders Society–revised Unified Parkinson's Disease Rating Scale; MoCA = Montreal Cognitive Assessment; PDBP = the Parkinson Disease Biomarkers Program; PIGD = postural instability and gait disorder.

**Supplementary Table 6. 2-year follow-up clinical characteristics by subtypes in the PDBP cohort**

| **Variables** | **Subtype PD-I (Inching Pace)** | **Subtype PD-M (Moderate Pace)** | **Subtype PD-R (Rapid Pace)** | **P-value[a]** | **Post-hoc[b]** | **P-value adjusted[c]** |
|---|---|---|---|---|---|---|
| **# of participants** | **55** | **72** | **49** | **-** | **-** | **-** |
| ***Motor manifestations*** | | | | | | |
| MDS-UPDRS Part II, mean (SD) | 4.2 (3.6) | 6.5 (4.4) | 14.6 (8.5) | <0.001 | III vs. rest | <0.001 |
| MDS-UPDRS Part III, mean (SD) | 13.5 (5.8) | 17.1 (7.8) | 27.3 (12.4) | <0.001 | III vs. rest | <0.001 |
| H&Y Stage, mean (SD) | 1.9 (0.4) | 1.9 (0.3) | 2.0 (0.7) | 0.572 | - | 0.708 |
| Schwab and England score, mean (SD) | 93.3 (6) | 92.2 (11.7) | 81.2 (17.7) | <0.001 | III vs. rest | <0.001 |
| Tremor score, mean (SD) | 0.4 (0.3) | 0.5 (0.3) | 0.5 (0.3) | 0.095 | - | 0.123 |
| PIGD score, mean (SD) | 0.1 (0.2) | 0.2 (0.2) | 0.6 (0.7) | <0.001 | III vs. rest | <0.001 |
| Motor phenotype, N (%) | | | | | | |
| Tremor | 27 (49.1) | 52 (72.2) | 21 (42.9) | | | |
| Indeterminate | 10 (18.2) | 2 (2.8) | 2 (4.1) | 0.005 | - | - |
| PIGD | 8 (14.6) | 6 (8.3) | 19 (38.8) | | | |
| ***Non-motor manifestations*** | | | | | | |
| MDS-UPDRS Part I, mean (SD) | 4.9 (3.8) | 7.7 (5.3) | 11.4 (6.3) | <0.001 | All comparisons | <0.001 |
| Hallucination, mean (SD) | 0.04 (0.21) | 0.03 (0.18) | 0.24 (0.73) | 0.036 | II vs. III | 0.022 |
| Apathy, mean (SD) | 0.1 (0.3) | 0.1 (0.4) | 0.4 (0.7) | 0.002 | III vs. rest | 0.002 |
| Pain, mean (SD) | 0.6 (0.7) | 0.8 (0.9) | 1.3 (1.2) | 0.005 | I vs. III | 0.002 |
| Fatigue, mean (SD) | 0.6 (0.8) | 1.1 (1.1) | 1.2 (1.0) | 0.015 | I vs. II | 0.004 |
| Sleep, mean (SD) | | | | | | |
| Epworth sleepiness score | 5.2(3.4) | 5.6 (4.4) | 10.1 (4.3) | <0.001 | III vs. rest | <0.001 |
| REM sleep behavior disorder | 0.1 (0.9) | 0.05 (0.3) | 0.2 (0.8) | 0.656 | - | 0.642 |
| Cognitive function, mean (SD) | | | | | | |
| MoCA-language | 2.5 (0.7) | 2.4 (0.8) | 2.1 (0.9) | 0.053 | - | 0.097 |
| MoCA total score | 27.6 (2.1) | 26.1 (2.8) | 24.5 (5.4) | <0.001 | I vs. III | 0.014 |

[a] P-values were calculated using ANOVA (for continuous variables) and $\chi^2$ test (for categorical variables) where appropriate.
[b] Post-hoc analysis was performed using the Tukey HSD test when the ANOVA P-value < 0.05.
[c] ANCOVA was used to calculate p-values (for continuous variables) adjusting for age and sex.

Abbreviations: MDS-UPDRS = Movement Disorders Society–revised Unified Parkinson's Disease Rating Scale; MoCA = Montreal Cognitive Assessment; PIGD = postural instability and gait disorder.

**Supplementary Table 7. 4-year follow-up clinical characteristics by subtypes in the PDBP cohort**

| Variables | Subtype PD-I (Inching Pace) | Subtype PD-M (Moderate Pace) | Subtype PD-R (Rapid Pace) | P-value[a] | Post-hoc[b] | P-value adjusted[c] |
|---|---|---|---|---|---|---|
| **# of participants** | **55** | **72** | **49** | **-** | **-** | **-** |
| ***Motor manifestations*** | | | | | | |
| MDS-UPDRS Part II, mean (SD) | 2.7 (1.7) | 8 (4.0) | 16.2 (6.6) | <0.001 | All comparisons | <0.001 |
| MDS-UPDRS Part III, mean (SD) | 17.9 (7.8) | 21.8 (5.8) | 31.7 (9.9) | 0.002 | III vs. rest | 0.003 |
| H&Y Stage, mean (SD) | 2.0 (0.0) | 2.0 (0.0) | 2.4 (0.7) | 0.022 | III vs. rest | 0.019 |
| Schwab and England score, mean (SD) | 95 (5.3) | 90.8 (4.9) | 76.7 (18.7) | 0.002 | III vs. rest | 0.003 |
| Tremor score, mean (SD) | 0.4 (0.3) | 0.4 (0.4) | 0.5 (0.2) | 0.714 | - | 0.598 |
| PIGD score, mean (SD) | 0.1 (0.1) | 0.4 (0.3) | 1.1 (0.5) | <0.001 | III vs. rest | <0.001 |
| Motor phenotype, N (%) | | | | | | |
| Tremor | 8 (14.6) | 5 (7.0) | 2 (4.1) | 0.056 | - | - |
| Indeterminate | 1 (1.9) | 4 (5.6) | 1 (2.0) | | | |
| PIGD | 1 (1.9) | 4 (5.6) | 6 (12.2) | | | |
| ***Non-motor manifestations*** | | | | | | |
| MDS-UPDRS Part I, mean (SD) | 4.2 (3.0) | 8.4 (2.9) | 13.6 (7.3) | <0.001 | III vs. rest | <0.001 |
| Hallucination, mean (SD) | - | - | 0.6 (0.7) | - | - | - |
| Apathy, mean (SD) | 0.2 (0.4) | 0.1 (0.3) | 0.3 (0.5) | 0.337 | - | 0.288 |
| Pain, mean (SD) | 0.5 (0.5) | 1.4 (1.3) | 1.6 (1.1) | 0.070 | - | 0.09 |
| Fatigue, mean (SD) | 0.6 (0.7) | 1.1 (1.0) | 1.2 (1.0) | 0.282 | - | 0.292 |
| Sleep, mean (SD) | | | | | | |
| Epworth sleepiness score | 4.4 (2.2) | 5.8 (3.1) | 9.9 (3.5) | 0.001 | III vs. rest | <0.001 |
| REM sleep behavior disorder | - | - | - | - | - | - |
| Cognitive function, mean (SD) | | | | | | |
| MoCA-language | 2.8 (0.4) | 2.5 (0.9) | 2.1 (1.1) | 0.209 | - | 0.309 |
| MoCA total score | 28.2 (1.9) | 24.4 (4.4) | 24.9 (3.6) | 0.039 | I vs. II | 0.096 |

[a] P-values were calculated using ANOVA (for continuous variables) and $\chi^2$ test (for categorical variables) where appropriate.
[b] Post-hoc analysis was performed using the Tukey HSD test when the ANOVA P-value < 0.05.
[c] ANCOVA was used to calculate p-values (for continuous variables) adjusting for age and sex.

Abbreviations: MDS-UPDRS = Movement Disorders Society–revised Unified Parkinson's Disease Rating Scale; MoCA = Montreal Cognitive Assessment; PIGD = postural instability and gait disorder.

**Supplementary Table 8. Annual progression rates in clinical manifestations and CSF biomarkers by subtypes assessed by linear mixed effects models in the PDB cohort**

| Variable | Subtype PD-I (Inching Pace) | | Subtype PD-M (Moderate Pace) | | Subtype PD-R (Rapid Pace) | |
|---|---|---|---|---|---|---|
| | ***β*** | **P value** | ***β*** | **P value** | ***β*** | **P value** |
| ***Motor manifestations*** | | | | | | |
| MDS-UPDRS Part II | 0.10 (-0.20, 0.40) | 0.530 | 1.01 (0.67, 1.36) | <0.001 | 2.53 (1.40, 3.67) | <0.001 |
| MDS-UPDRS Part III | -0.30 (-1.07, 0.47) | 0.444 | 1.12 (0.46, 1.78) | 0.002 | 2.70 (1.70, 3.70) | <0.001 |
| H&Y Stage | 0.04 (-0.00, 0.08) | 0.078 | 0.05 (0.02, 0.08) | 0.007 | 0.10 (0.01, 0.19) | 0.042 |
| Schwab and England score | -0.48 (-1.10, 0.11) | 0.117 | -0.28 (-1.10, 0.53) | 0.505 | -4.05 (-5.92, -2.18) | <0.001 |
| Tremor score | -0.01 (-0.03, 0.02) | 0.706 | 0.01 (-0.02, 0.03) | 0.465 | 0.03 (-0.00, 0.06) | 0.095 |
| PIGD score | -0.03 (-0.04, -0.01) | <0.001 | 0.02 (-0.00, 0.041) | 0.110 | 0.14 (0.06, 0.22) | 0.001 |
| ***Non-motor manifestations*** | | | | | | |
| MDS-UPDRS Part I | 0.11 (-0.10, 0.32) | 0.333 | 0.85 (0.53, 1.16) | <0.001 | 1.36 (0.91, 1.82) | <0.001 |
| Hallucination | 0.01 (-0.01, 0.03) | 0.422 | 0.01 (-0.01, 0.02) | 0.377 | 0.06 (0.01, 0.12) | 0.031 |
| Apathy | -0.00 (-0.05, 0.04) | 0.853 | 0.04 (-0.01, 0.09) | 0.162 | 0.05 (-0.02, 0.13) | 0.187 |
| Pain | -0.03 (-0.09, 0.04) | 0.453 | 0.13 (0.07, 0.20) | <0.001 | 0.14 (0.03, 0.24) | 0.013 |
| Fatigue | -0.02 (-0.08, 0.03) | 0.440 | 0.09 (0.02, 0.16) | 0.023 | 0.13 (0.06, 0.20) | 0.002 |
| ***Sleep*** | | | | | | |
| Epworth sleepiness score | 0.12 (-0.11, 0.34) | 0.327 | 0.17 (-0.14, 0.47) | 0.294 | 0.69 (0.23, 1.10) | 0.005 |
| REM sleep behavior disorder[a] | - | - | - | - | - | - |
| ***Cognitive function*** | | | | | | |
| MoCA-language | 0.03 (-0.05, 0.11) | 0.469 | 0.01 (-0.07, 0.08) | 0.845 | -0.16 (-0.26, -0.06) | 0.005 |
| MoCA total score | 0.02 (-0.15, 0.18) | 0.836 | -0.15 (-0.4, 0.09) | 0.214 | -1.10 (-1.62, -0.57) | <0.001 |

[a] The values in REM sleep behavior disorders are so sparse that the corresponding beta is not available.
Abbreviations: MDS-UPDRS = Movement Disorders Society–revised Unified Parkinson's Disease Rating Scale; MoCA = Montreal Cognitive Assessment; PDBP = the Parkinson Disease Biomarkers Program; PIGD = postural instability and gait disorder.

**Supplementary Table 9. Baseline CSF biomarkers by subtypes in the PPMI cohort**

| Biomarker | HC | PD-I, mean (SD) | PD-M, mean (SD) | PD-R, mean (SD) | P values[a] | | | | | |
|---|---|---|---|---|---|---|---|---|---|---|
| | | | | | HC vs. PD-I | HC vs. PD-M | HC vs. PD-R | PD-M vs. PD-I | PD-R vs. PD-I | PD-R vs. PD-M |
| $\alpha$-synuclein | 1704.491 (752.640) | 1607.109 (734.891) | 1487.264 (659.634) | 1357.906 (505.550) | 0.020 | 0.001 | 0.218 | 0.153 | 0.011 | 0.068 |
| A$\beta$-42 | 1025.042 (498.628) | 970.128 (463.830) | 905.070 (386.737) | 781.500 (351.559) | 0.148 | 0.398 | 0.912 | 0.227 | 0.009 | 0.045 |
| P-tau | 16.845 (8.412) | 14.342 (5.403) | 14.014 (5.107) | 14.959 (6.352) | 0.013 | <0.001 | 0.023 | 0.571 | 0.767 | 0.760 |
| T-tau | 190.283 (79.901) | 169.613 (59.369) | 163.533 (53.990) | 170.126 (66.713) | 0.014 | <0.001 | 0.017 | 0.319 | 0.437 | 0.933 |
| A$\beta$-42/T-tau | 5.578 (1.649) | 5.713 (1.480) | 5.635 (1.572) | 4.906 (1.885) | 0.479 | <0.001 | 0.009 | 0.962 | 0.022 | 0.051 |
| A$\beta$-42/$\alpha$-synuclein | 0.636 (0.221) | 0.637 (0.219) | 0.659 (0.282) | 0.593 (0.208) | 0.182 | <0.001 | 0.218 | 0.311 | 0.340 | 0.402 |
| P-tau/$\alpha$-synuclein | 0.010 (0.002) | 0.009 (0.002) | 0.010 (0.002) | 0.011 (0.003) | 0.821 | 0.742 | 0.004 | 0.072 | 0.001 | 0.007 |
| P-tau/T-tau | 0.087 (0.007) | 0.084 (0.008) | 0.085 (0.007) | 0.087 (0.008) | 0.057 | 0.228 | 0.140 | 0.215 | 0.114 | 0.145 |
| T-tau/$\alpha$-synuclein | 0.116 (0.026) | 0.113 (0.028) | 0.117 (0.029) | 0.128 (0.026) | 0.664 | 0.400 | 0.017 | 0.219 | 0.015 | 0.036 |
| A$\beta$-42/P-tau | 64.795 (20.617) | 68.010 (17.636) | 66.679 (20.169) | 56.961 (22.692) | 0.207 | <0.001 | 0.006 | 0.841 | 0.010 | 0.037 |

[a]ANCOVA was used to calculate p-values adjusting for age and sex.

Abbreviations: A$\beta$-42 = the 42 amino acid form of amyloid-$\beta$; CSF = cerebrospinal fluid.
